# Minimally invasive 1 mm skin biopsies capture regional transcriptomic heterogeneity in vitiligo

**DOI:** 10.1101/2025.11.21.25340665

**Authors:** Danique Berrevoet, Arno Belpaire, Elise Van Caelenberg, Barbara Boone, Guillaume Cattebeke, Yannick Gansemans, Elise Callens, Koen Deserranno, Dieter Deforce, Reinhart Speeckaert, Filip Van Nieuwerburgh

## Abstract

Vitiligo is an autoimmune depigmentation disorder characterized by melanocyte loss and complex immune dysregulation. Despite therapeutic advances, variable efficacy and frequent relapse highlight the need for deeper insight into its pathophysiology. Conventional 3–5 mm skin biopsies are invasive, induce visible scars and require suturing, limiting their use in clinical and translational research. Here, we demonstrate that minimally invasive 1 mm punch biopsies yield adequate RNA for bulk transcriptomic profiling, enabling comprehensive molecular characterization of vitiligo skin. RNA sequencing of 105 biopsies spanning lesional, perilesional, non-lesional, and control skin revealed distinct region-specific transcriptional signatures involving interferon signaling, immune activation, and metabolic reprogramming. Beyond canonical melanocyte and immune pathways, we identify underexplored dysregulation of cellular clearance and structural regulatory mechanisms, implicating ferroptosis, peroxisome, and efferocytosis pathways in lesion persistence. Connectivity Map analysis further predicted candidate compounds capable of reversing the lesional transcriptional signature, pointing to epigenetic regulators, tyrosine kinase inhibitors, and metabolic modulators. These findings establish 1 mm skin biopsies as a feasible and biologically informative tool for minimally invasive molecular profiling and highlight new therapeutic axes in vitiligo pathogenesis.

## Introduction

Vitiligo is the most common skin depigmentation disorder worldwide and is characterized by selective loss of melanocytes, resulting in depigmented lesions (1). Substantial progress has been made in elucidating its pathogenesis, leading to its classification as an autoimmune disorder. Central to this understanding is the identification of immunological mechanisms in which melanocyte-specific cytotoxic T cells mediate targeted melanocyte destruction, a process in which the Janus Kinase/Signal Transducer and Activator of Transcription (JAK-STAT) signaling pathway plays a critical role (2). While these insights have facilitated the development of promising targeted therapies such as Janus kinase (JAK) inhibitors, currently available treatments are still limited by side effects, inconsistent efficacy, and high relapse rates upon discontinuation (2–4). This variability in treatment response may be partly attributed to the involvement of alternative or additional immune pathways beyond the well-characterized JAK-STAT signaling axis, operating within the local skin microenvironment. Defining these lesion-resident pathways can inform mechanism-based treatments, highlighting agents predicted to correct disease-associated states.

Transcriptomic profiling of skin offers a high-resolution approach to uncover local immune and metabolic changes, yet its routine application in dermatology is constrained by the invasiveness of standard 3–5 mm punch biopsies, which induce scars, require suturing, and are impractical for repeated sampling or routine clinical use (5,6). While other non-invasive approaches such as tape-stripping have been explored, they are limited to the superficial layers of the epidermis and fail to capture the full cellular complexity of the skin microenvironment (7). Moreover, limited studies have systematically compared lesional, perilesional, and non-lesional skin from the same vitiligo patient using transcriptomic approaches, limiting our understanding of disease heterogeneity within individuals. Emerging evidence suggests that vitiligo extends beyond visibly depigmented lesions. Perilesional skin, representing the transition between lesional and unaffected areas, exhibits active immune infiltration, while non-lesional skin may harbor subclinical inflammatory or metabolic alterations indicative of a prelesional state (8–10). This spatial heterogeneity highlights the importance of region-specific molecular profiling to delineate the continuum of disease activity within individual patients.

In this study, we apply bulk RNA sequencing to 1 mm punch biopsies from lesional, perilesional, non-lesional, and control skin to evaluate the feasibility and biological fidelity of minimally invasive transcriptomics. Our approach aims to define region-specific transcriptional signatures that capture both established and previously underrecognized disease pathways and to explore candidate pharmacologic modulators capable of reversing the vitiligo-associated expression profile. Through this framework, we seek to demonstrate the value of small-biopsy transcriptomics as a practical and scalable tool for molecular discovery and translational research in vitiligo and other skin diseases.

## Materials & Methods

### Patient inclusion

A total of 28 patients with non-segmental vitiligo were recruited between July 2024 and May 2025 at the Department of Dermatology, Ghent University Hospital. The study was approved by the Ethics Committee of University Hospital Ghent (Belgium), and all participants provided written informed consent prior to enrollment. Inclusion criteria comprised a confirmed diagnosis of non-segmental vitiligo, age ≥18 years, and the ability to provide informed consent. Individuals with segmental vitiligo or concurrent immune-mediated inflammatory disorders were excluded. A total of 27 healthy control subjects were also recruited. Clinical and demographic data collected for each participant included age, sex, BMI, age at disease onset, family history, current treatment, smoking status, disease status, and disease activity. Disease activity was clinically assessed using standardized follow-up photographs and quantified with the Vitiligo Disease Activity Score (VDAS) and Vitiligo Disease Improvement Score (VDIS) (11). Patients were classified as having active disease (disease status: 1) if objectively evaluated before-and-after photographs demonstrated an increase in VDAS score, typically assessed over a 6-month interval up to a maximum of 12 months. In the absence of photographic documentation (e.g., at the initial consultation), active disease status was determined based on a self-reported mVIDA score indicating disease activity within the preceding 3 months. Demographic and clinical characteristics of all participants are summarized in Table 1.

**Table 1.**
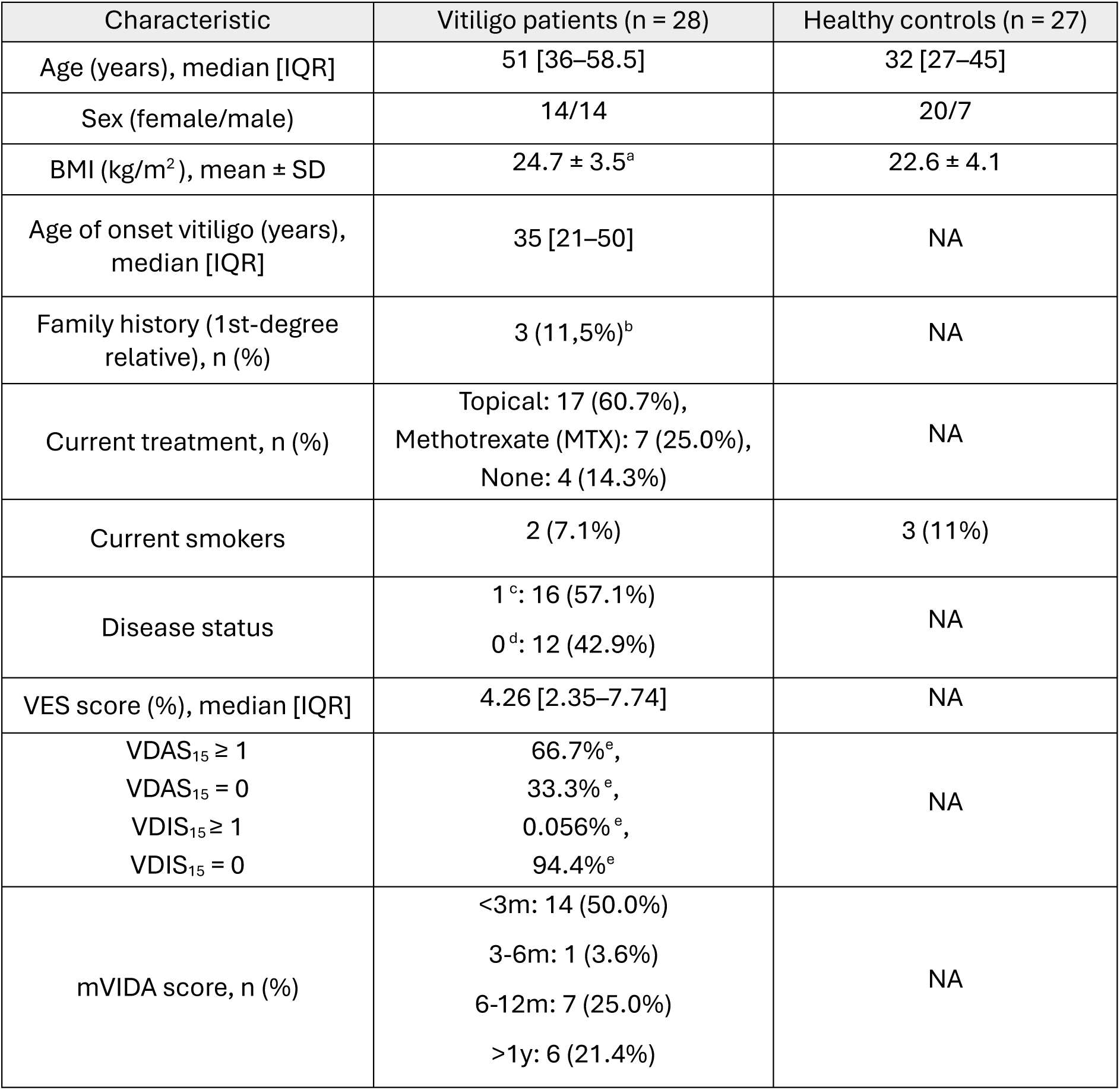
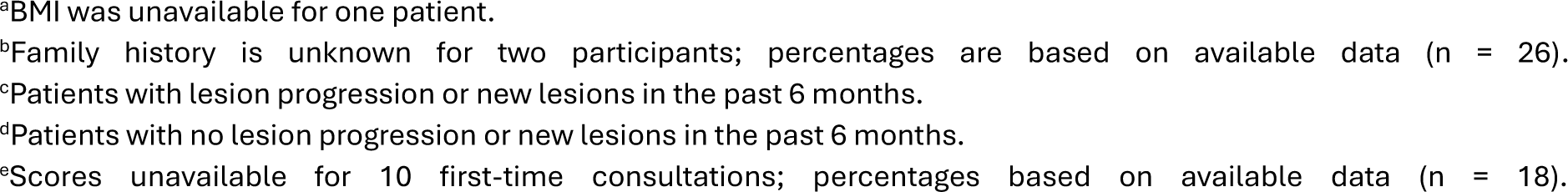
Participant characteristics.

| Characteristic | Vitiligo patients ($n = 28$) | Healthy controls ($n = 27$) |
| --- | --- | --- |
| Age (years), median [IQR] | 51 [36–58.5] | 32 [27–45] |
| Sex (female/male) | 14/14 | 20/7 |
| BMI ($\text{kg}/\text{m}^2$), mean ± SD | 24.7 ± 3.5<sup>a</sup> | 22.6 ± 4.1 |
| Age of onset vitiligo (years), median [IQR] | 35 [21–50] | NA |
| Family history (1st-degree relative), $n$ (%) | 3 (11.5%)<sup>b</sup> | NA |
| Current treatment, $n$ (%) | Topical: 17 (60.7%),<br>Methotrexate (MTX): 7 (25.0%),<br>None: 4 (14.3%) | NA |
| Current smokers | 2 (7.1%) | 3 (11%) |
| Disease status | 1<sup>c</sup>: 16 (57.1%)<br>0<sup>d</sup>: 12 (42.9%) | NA |
| VES score (%), median [IQR] | 4.26 [2.35–7.74] | NA |
| VDAS<sub>15</sub> ≥ 1<br>VDAS<sub>15</sub> = 0<br>VDIS<sub>15</sub> ≥ 1<br>VDIS<sub>15</sub> = 0 | 66.7%<sup>e</sup>,<br>33.3%<sup>e</sup>,<br>0.056%<sup>e</sup>,<br>94.4%<sup>e</sup> | NA |
| mVIDA score, $n$ (%) | <3m: 14 (50.0%)<br>3–6m: 1 (3.6%)<br>6–12m: 7 (25.0%)<br>>1y: 6 (21.4%) | NA |

<sup>a</sup>BMI was unavailable for one patient.
<sup>b</sup>Family history is unknown for two participants; percentages are based on available data (n = 26).
<sup>c</sup>Patients with lesion progression or new lesions in the past 6 months.
<sup>d</sup>Patients with no lesion progression or new lesions in the past 6 months.
<sup>e</sup>Scores unavailable for 10 first-time consultations; percentages based on available data (n = 18).

### Sample collection

Skin punch biopsies with a diameter of 1 mm were obtained under local anaesthesia (xylocaine 2% + adrenaline) using sterile punch tools (Kai Europe GmbH, Germany). Where feasible, each patient contributed lesional (depigmented), perilesional (lesion border), and non-lesional skin (clinically unaffected, ∼1 cm from the border) to minimize inter-individual variability. In three cases, only one or two biopsies were obtained due to anatomical or patient-related constraints. In total, 24 lesional, 27 perilesional and 27 non-lesional skin biopsies were collected from 28 patients. The punch biopsies were preferentially taken from the arm or leg to ensure consistency in anatomical location. For the external healthy control group (n = 27), one biopsy per individual was collected from the upper arm. Immediately after excision, all biopsies were snap frozen in liquid nitrogen and stored at −80°C until further processing.

### Sample processing

Immediately upon retrieval from −80 °C, 0.3 mL of TriReagent (Zymo Research, Irvine, CA, USA) was added to each 1 mm skin punch biopsy. To minimise RNA degradation, SuperaseIn RNase Inhibitor (Thermo Fisher Scientific, Waltham, MA, USA) was added. The tissue was lysed mechanically using a 1000 µL pipette tip, followed by shearing through a syringe (Terumo Corporation, Tokyo, Japan) fitted with a 20Gx1½ needle (Terumo Corporation). Total RNA was extracted using the conventional TriReagent/chloroform method and subsequently purified with the Direct-zol Microprep RNA Kit (Zymo Research), including on-column DNase I treatment, according to the manufacturer’s instructions. As residual genomic DNA was detected in some samples, an additional DNase treatment was performed using the TURBO DNA-*free*™ Kit (Invitrogen, Thermo Fisher Scientific) on all RNA extracts. RNA concentration was measured using the Qubit® 2.0 Fluorometer with Qubit RNA HS Assay Kit (Thermo Fisher Scientific), and RNA integrity was assessed via capillary electrophoresis using the 5200 Fragment Analyzer (Agilent Technologies, Santa Clara, CA, USA). Libraries were prepared using the QuantSeq 3’ mRNA-Seq V2 Library Prep Kit FWD with UDI (Lexogen, Vienna, Austria), starting from 28 ng total RNA per sample. Biopsies from healthy controls (n = 27) were processed using the same protocol. RNA quantity and integrity from the 1 mm samples consistently met the requirements for library preparation.

### RNA sequencing and preprocessing

Bulk RNA sequencing of all 105 samples was performed on the AVITI™ Cloudbreak Freestyle platform (high-output kit), generating 75 bp single-end reads. Raw sequencing reads were assessed for quality and length using FastQC (v0.12.1)(12). Putative contaminations were evaluated with FastQ Screen (v0.15.3) against a panel of genomes from common laboratory organisms. Adapter trimming was performed using cutadapt (v5.0), with additional filtering to remove reads containing ambiguities or failing to meet a minimum phred quality score of 20 (13). The quality of the trimmed read pairs was re-evaluated using FastQC. Sequence alignment was performed using STAR aligner (v2.7.11b) against the human reference genome (GRCh38, ENSEMBL release 112) (14). UMI-based deduplication of mapped reads was performed with UMI-tools (v1.1.6), and feature counting at the gene and transcript isoform level was done using rsem-calculate-expression (RSEM; v1.3.1)(15,16). The gene-level counts were used for all downstream analyses in R (v4.5.0).

### Protein-coding gene detection and clustering analysis

Gene annotations were imported from the reference GTF via rtracklayer (v1.68.0) and integrated with the obtained counts using dplyr (v1.1.4) and tidyr (v1.3.1). To quantify detected protein-coding genes, only entries annotated as protein_coding in the GTF were retained. For each sample, detected genes (counts > 0) were tallied per biotype, and biotype composition was expressed as the percentage of detected genes belonging to each biotype. Principal component analysis (PCA) was performed on the rlog-transformed (DESeq2; v1.48.1) counts using prcomp to assess sample clustering and batch effects across all samples.

### Differential expression analysis

Differential gene expression analysis was conducted using EdgeR (v4.2.2) (17). Feature counts were TMM-normalized and correction of the P values for repeated testing (adjusted P value (P_adj_)) was performed using the Benjamini-Hochberg method. Genes with P_adj_ < 0.05 and |log_2_FC|>=1 (≥2-fold) were considered significantly differentially expressed. Samples were classified as lesional, perilesional, non-lesional, or control. Tested contrasts: lesional vs control, perilesional vs control, non-lesional vs control, lesional vs perilesional, lesional vs non-lesional, and perilesional vs non-lesional. Results are reported as Group 1 vs Group 2, with positive log₂FC indicating higher expression in Group 1. Gene symbols follow HGNC nomenclature.

### Gene Set Enrichment Analysis

Gene Set Enrichment Analysis (GSEA) was performed using clusterprofiler on the full ranked (logFC) gene lists for each contrast (18). Analyses were conducted using annotated gene sets from the Kyoto Encyclopedia of Genes and Genomes (KEGG; release 116.0) and Gene Ontology (GO; v3.21.0) databases. Enriched pathways were considered significant if P_adj_ < 0.05.

### Connectivity Map analysis

Connectivity Map (CMap) queries were run on the CLUE platform (with BING extension) using the lesional vs control DEG set. Genes were ranked by a composite of log₂FC and P_adj_ and split into up- and down-regulated lists. Only DEGs with |log₂FC| ≥ 0.585 (≥1.5-fold) and P_adj_ < 0.05 were eligible. As CMap queries accept ≤150 genes per direction, genes outside the L1000+BING universe were replaced by the next highest-ranked DEGs (ranked by |log₂FC| × −log₁₀ (P_adj_), and if >150 valid genes were available, the top 150 were used. Analyses were restricted to small-molecule perturbations (trt_cp) in the A375 (melanocytic) cell line. Similarity was scored by the raw connectivity score (raw_cs) (positive = similar; negative = inverse). Significance used fdr_q_nlog10 and high-confidence hits were defined as fdr_q_nlog10 ≥ 15.35 (q < 10⁻¹⁵.³⁵).

## Results

### Participant characteristics and study overview

We conducted a cross-sectional study using 105 1 mm skin punch biopsies from 28 patients with non-segmental vitiligo and 27 healthy controls. Participant characteristics are summarized in Table 1. Each patient contributed lesional (n = 24), perilesional (n = 27), and non-lesional (n = 27) samples when feasible. All biopsies met the minimum RNA input requirement (10 ng total RNA per sample) suitable for library preparation, confirming the feasibility of minimally invasive transcriptomics. The cohort represented a balanced sex distribution (50% female) with a median age of 51 years (IQR 36–59) and median disease onset at 35 years. Sixteen patients (57%) exhibited clinically active disease based on standardized VDAS/VDIS and mVIDA assessments. An overview of the experimental design and analysis workflow is shown in Figure 1.

**Figure 1.**
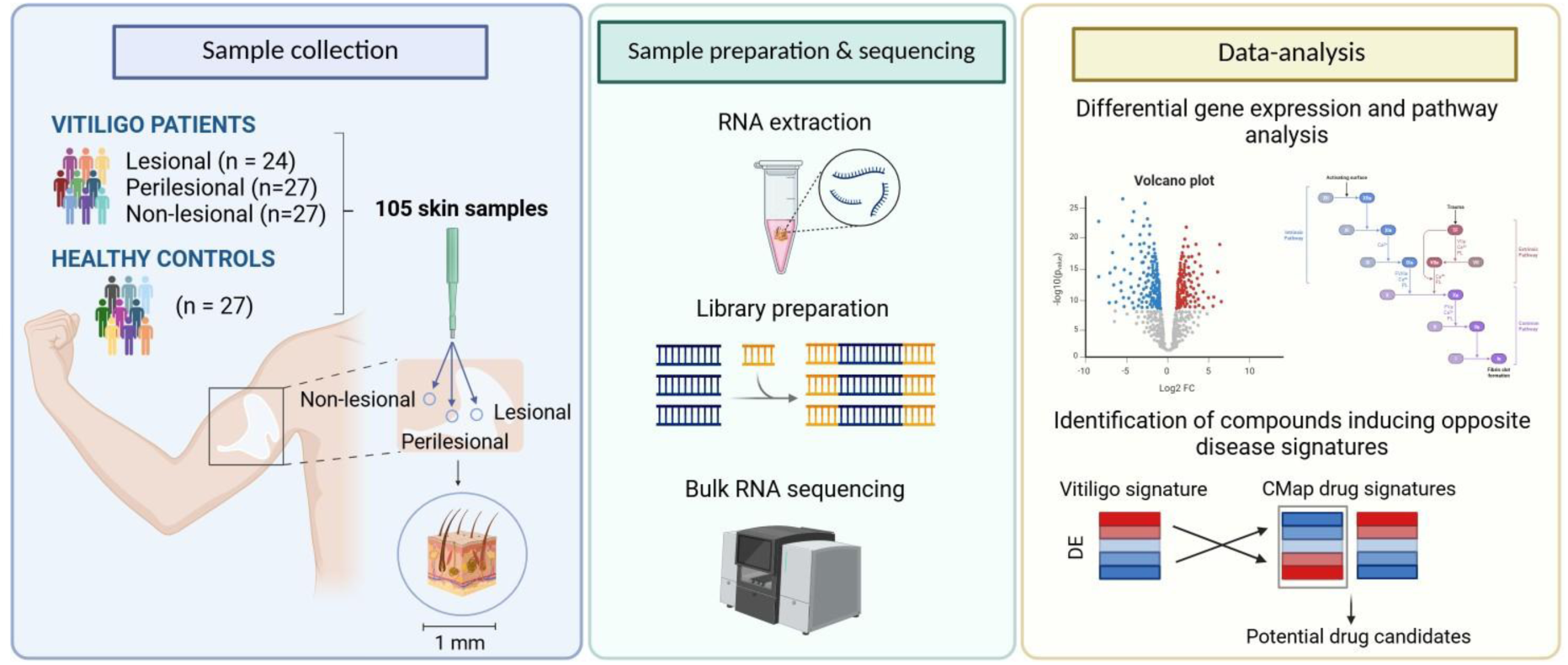
Overview of the study design. The schematic shows 1 mm skin punch biopsy sampling from lesional, perilesional, non-lesional, and control skin, followed by bulk RNA sequencing and integrative bioinformatic analyses. Downstream analyses included differential expression, pathway enrichment, and CMap–based identification of compounds predicted to reverse vitiligo-associated transcriptional signatures. DE: differential expression.

### Global transcriptional characterization of vitiligo skin

Bulk RNA sequencing achieved a mean of 38.7 million reads per sample, detecting approximately 15,000 protein-coding genes per biopsy (Figure S1). PCA revealed distinct separation between lesional and control skin, reflecting extensive transcriptional reprogramming in established vitiligo lesions (Figure 2A). Perilesional and non-lesional samples showed substantial overlap and partial proximity to lesional skin, suggestive of early or subclinical disease-related activity beyond visibly affected areas.

**Figure 2.**
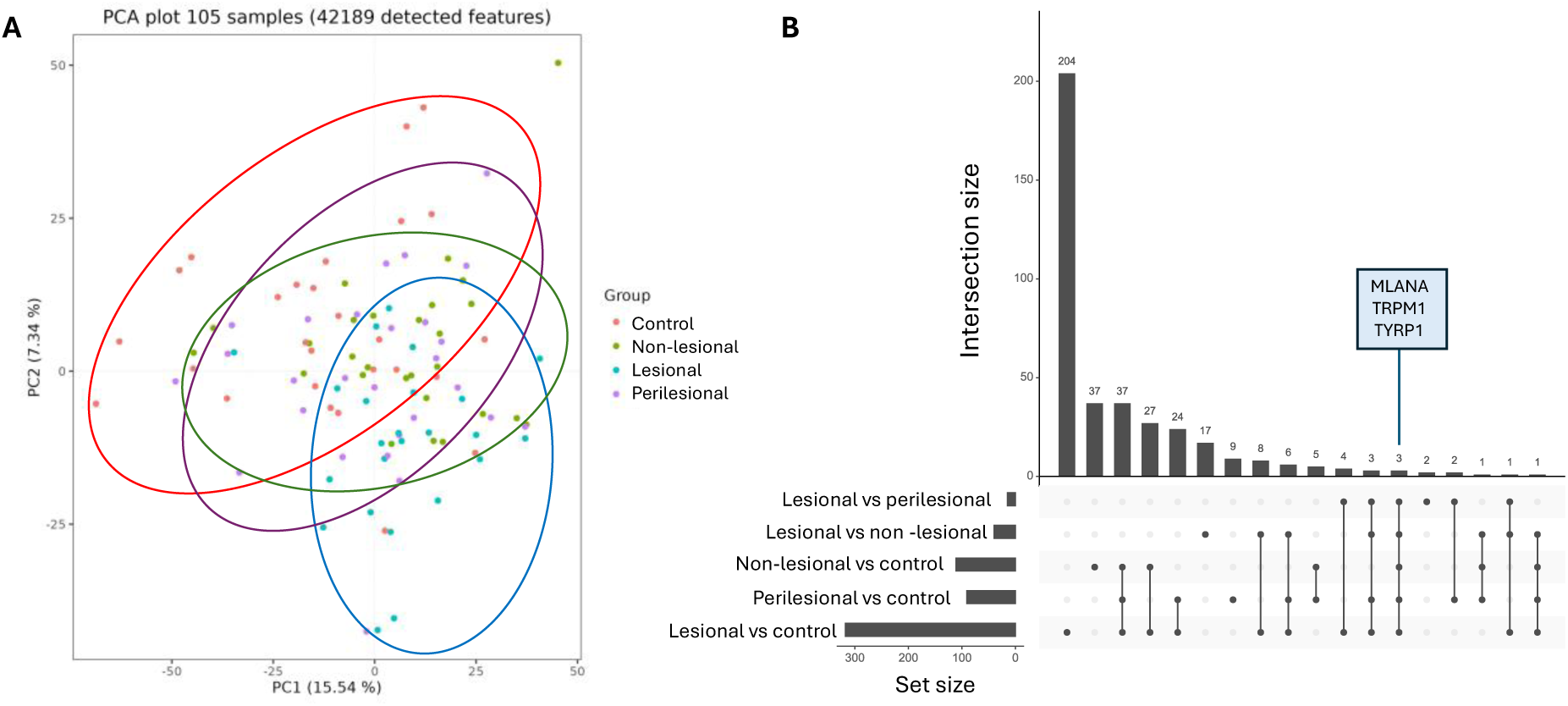
A) PCA of transcriptomic profiles from lesional, perilesional and non-lesional skin from vitiligo patients, and skin samples from healthy controls. B) UpSet plot showing overlapping DEGs across comparisons. The set size represents the total number of DEGs identified in each individual comparison, while the intersection size indicates the number of DEGs shared between different comparisons, as indicated by the connected dots below the bars. This visualization highlights both unique and shared transcriptional changes across skin regions in vitiligo. The perilesional versus non-lesional comparison is excluded, as no significant DEGs were detected in this comparison (|log2FC|>=1, Padj < 0.05).

Differential expression analysis (Figure S2, panels A-F) identified 320 DEGs in lesional versus control skin (45 upregulated, 275 downregulated), 92 DEGs in perilesional versus control skin (30 upregulated, 62 downregulated), and 124 DEGs in non-lesional versus control skin (63 upregulated, 61 downregulated). Within-patient comparisons revealed 40 DEGs between lesional and non-lesional skin (4 upregulated, 36 downregulated), 15 DEGs (all upregulated) between perilesional and lesional skin, and no significant changes between perilesional and non-lesional skin (Table S1). The majority of DEGs were unique to the lesional versus control comparison, reflecting the distinct transcriptional profile of lesional vitiligo skin (Figure 2B). Only 3 genes (*MLANA, TRPM1, TYRP1*) were common to all comparisons.

### Minimally invasive transcriptomics captures canonical, novel and low-expression vitiligo signatures

The top differentially expressed genes across all comparisons reflected established features of vitiligo pathogenesis (Figure 3A). Melanocyte- and pigmentation-associated transcripts (e.g. *TYR, PMEL, DCT, PCSK2*) were markedly downregulated in lesional skin, reflecting melanocyte depletion in depigmented areas. Additionally, perilesional skin exhibited upregulation of inflammation-associated transcripts including *IFI44*, *IFI6*, and *FOSB*, consistent with active interferon signaling and early immune activation at sites of ongoing pigment loss.

**Figure 3.**
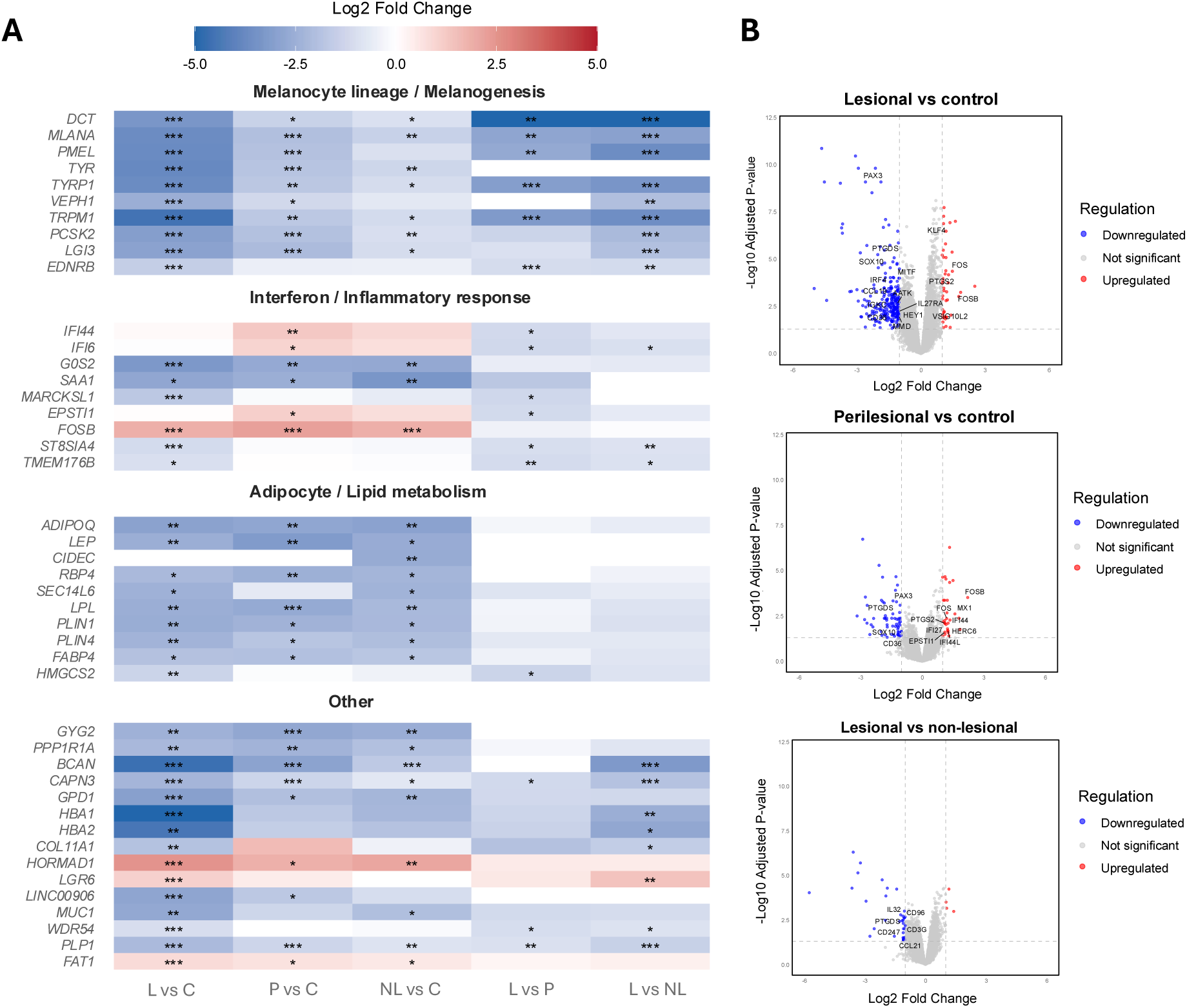
A) Combined set of top 15 DEGs across pairwise comparisons. The perilesional versus non-lesional comparison was excluded due to the absence of significant DEGs. Genes are displayed on the y-axis and grouped by functional category, based on their primary biological roles as defined in the literature. Cell color indicates the log2 fold change (blue: downregulation; red: upregulation), with statistical significance indicated by asterisks (*Padj* < 0.05, \**; < 0.01, **; < 0.001, \*\*\**). L, lesional; P, perilesional; NL, non-lesional; C, control. B) Volcano plots highlight immune-related transcriptional changes in vitiligo skin. Volcano plots display DEGs in lesional versus control, perilesional versus control, and lesional versus non-lesional skin. Immune-related genes including cytokines, chemokines, interferon-stimulated genes, transcription factors, and immune modulators, are annotated and color-coded (blue: downregulation; red: upregulation, grey: not significant).

Beyond canonical melanocytic and immune genes, several underexplored candidates (e.g. *PPP1R1A, FAT1, HORMAD1*) emerged among the most dysregulated transcripts, suggesting additional signaling and regulatory processes not previously implicated in vitiligo. The dataset also captured low abundant but biologically meaningful immune transcripts, including cytokine and chemokine signaling molecules (*IL27RA, IL32, IL4I1, CCL19, CCL21*), immune modulators (*CD36, CD96, CD3G, CD247*), transcription factors (*KLF4, PAX3, IRF4, MITF, FOS, FOSB, HEY1, SOX10*), and a strong type I interferon response signature (*IFI44, IFI27, IFI44L, IFI6, MX1, EPSTI1, HERC6*) (Figure 3B). Three comparisons are shown in Figure 3B; the remaining data are provided in Figure S3. Collectively, these results confirm that 1 mm biopsies yield sufficient depth and sensitivity to resolve both dominant and subtle molecular features of vitiligo skin.

### Pathway-level dysregulation across skin regions

GSEA revealed consistent negative enrichment (Normalized Enrichment Score (NES) < 0; P_adj_ < 0.05) of metabolic pathways in vitiligo skin, including oxidative phosphorylation, peroxisome proliferator-activated receptor (PPAR) signaling, and fatty acid metabolism (Figure 4). In parallel, immune and inflammatory programs were enriched (NES > 0; P_adj_ < 0.05) in perilesional and non-lesional skin, encompassing significant signals in tumor necrosis factor (TNF), NOD-like receptor (NLR) and T helper (Th)1/Th2 cell differentiation pathways, as well as antigen processing and presentation. Several less-studied pathways, such as phagosome, peroxisome, ferroptosis, and efferocytosis, were also significantly altered, indicating additional layers of cellular dysregulation. Additionally, “negative regulation of gene expression, epigenetic” (GO:0045814) was positively enriched in lesional versus control skin (Table S2), hinting at altered chromatin-mediated transcriptional control and supporting the therapeutic relevance of epigenetic modulators identified later by CMap analysis. Full GSEA results are available in Supplementary Table S3.

**Figure 4.**
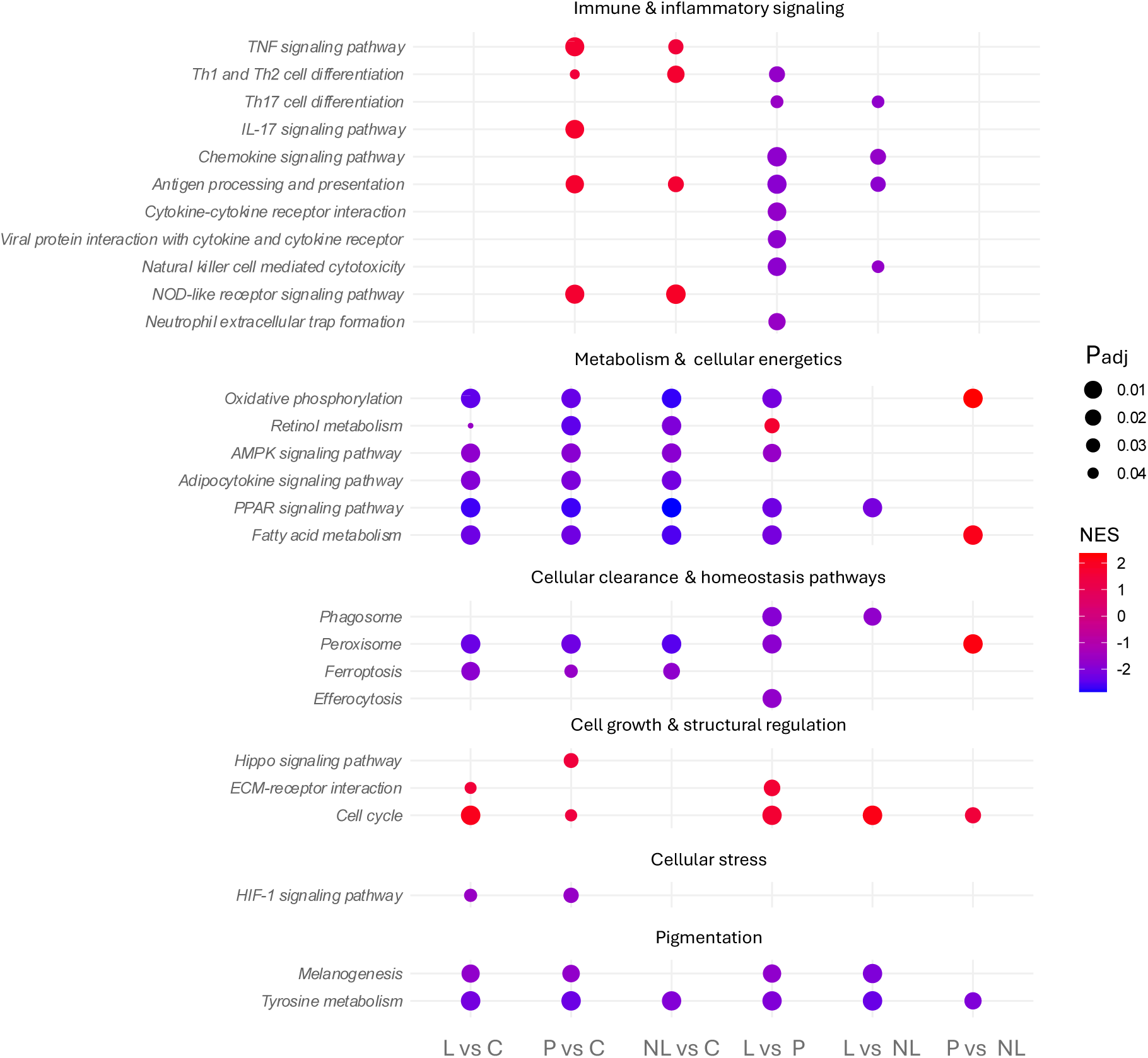
Dot plot summarizing KEGG pathways of interest significantly enriched across pairwise comparisons. Each dot denotes one KEGG pathway; the x-axis shows the indicated comparisons and the y-axis lists pathways. Dot color encodes NES (red = positive enrichment, blue = negative enrichment). Dot size reflects significance and is inversely proportional to the Padj (larger dots indicate smaller Padj). L, lesional skin; P, perilesional skin; NL, non-lesional skin; C, control skin.

### Connectivity Map analysis identifies candidate signature-reversing compounds

CMap analysis of the ranked lesional versus control gene expression signature (|log₂FC| ≥ 0.585) revealed compounds inducing inverse transcriptional profiles relative to vitiligo lesions. After applying filtering criteria (trt_cp; FDR –log₁₀ ≥ 15.35; A375 cell line), 170 high-confidence candidates were retained (Table S4) and subsequently screened for previously reported or mechanistically plausible links to vitiligo. Established vitiligo treatments such as JAK2 inhibitor NVP-BSK805 (raw_cs = −0.38), glucocorticoids (dexamethasone (raw_cs = −0.38), hydrocortisone (raw_cs = −0.39), medrysone (raw_cs = −0.35)), and methotrexate (raw_cs = −0.35) were identified among the negatively correlated signatures, validating the biological relevance of the dataset.

Beyond these reference compounds, epigenetic regulators targeting histone methylation (LL - 507, UNC-1999, BIX-01294, and GSK-343) were among the strongest inverse hits, consistent with the positive enrichment of epigenetic silencing pathways observed in lesional skin (Table S2). Inhibitors of growth factor and tyrosine kinase signaling (golvatinib, cediranib, lapatinib, ARR - 334543, CGP-53353, dasatinib, SU-11274) also showed strong negative connectivity, indicating that modulation of (vascular) epidermal growth factor (VEGFR/EGFR) and related non-JAK tyrosine kinase pathways may counteract disease-associated transcriptional states. In addition, immune and metabolism-linked modulators such as STA-5326, ibrutinib, PRT-062607, dipyridamole and PF-543, also demonstrated inverse transcriptional relationships, reflecting perturbation of their respective pathways; cytokine, Bruton’s tyrosine kinase (BTK), spleen tyrosine kinase (S K), phosphodiesterase (PDE) and sphingosine kinase signaling (SphK). Collectively, these findings highlight multiple druggable entry points converging on chromatin remodeling, immune regulation, and cellular metabolism (Table 2).

**Table 2.** Candidate drug repurposing compounds predicted to reverse the vitiligo transcriptional signature ranked by correlation score.

| Cell line | Perturbagen name | Mode of action | Connectivity score | FDR ($-\log_{10}$)<sup>a</sup> |
| --- | --- | --- | --- | --- |
| A375 | LLY-507 | SMYD2 inhibitor | -0.44 | *** |
| A375 | Cediranib | VEGFR inhibitor | -0.44 | *** |
| A375 | Golvatinib | VEGFR inhibitor | -0.43 | *** |
| A375 | SU-11274 | Tyrosine kinase inhibitor | -0.42 | *** |
| A375 | UNC-1999 | Histone lysine methyltransferase inhibitor | -0.40 | *** |
| A375 | dipyridamole | Phosphodiesterase inhibitor | -0.39 | *** |
| A375 | STA-5326 | Interleukin inhibitor | -0.37 | *** |
| A375 | ARRY-334543 | EGFR inhibitor | -0.35 | *** |
| A375 | CGP-53353 | EGFR inhibitor | -0.39 | *** |
| A375 | GSK-343 | Histone lysine methyltransferase inhibitor | -0.38 | *** |
| A375 | GSK-269962 | ROCK inhibitor | -0.38 | *** |
| A375 | BIX-01294 | Histone lysine methyltransferase inhibitor | -0.36 | *** |
| A375 | ibrutinib | BTK inhibitor | -0.35 | *** |
| A375 | lapatinib | EGFR inhibitor | -0.35 | *** |
| A375 | PRT-062607 | Syk inhibitor | -0.34 | *** |
| A375 | PF-543 | Sphingosine kinase inhibitor | -0.34 | *** |
| A375 | dasatinib | Tyrosine kinase inhibitor | -0.34 | *** |

<sup>a</sup>As per CMap conventions, FDR significance values are reported as $-\log_{10}(q)$ . Scores above 15.35 indicating extremely strong enrichment ( $q < 10^{-15.35}$ ) are reported in this table as '\*\*\*' ( $< 0.001$ ).

## Discussion

This study shows that 1 mm punch biopsies provide sufficient RNA yield and quality for comprehensive bulk transcriptomic profiling, enabling minimally invasive, suture-free molecular analysis in vitiligo. Despite their small size, they consistently produced high-coverage sequencing data and captured core disease-related transcriptional programs, supporting a low-input, high-output workflow. To our knowledge, this is the first application of 1 mm punch–based transcriptomics in vitiligo, extending a micro-biopsy approach that is gaining traction in dermatology (19). Their minimal invasiveness allows repeated longitudinal sampling and precise, site-directed molecular readouts. While smaller samples may underrepresent broader heterogeneity, they uniquely enable focused interrogation of local microenvironmental changes with high spatial specificity. These findings demonstrate the feasibility of 1 mm biopsy transcriptomics as a practical and patient-friendly platform for molecular dermatology and longitudinal immune monitoring.

At the biological level, the data delineate a spatial gradient of transcriptional changes across lesional, perilesional, and non-lesional skin. The expected downregulation of melanogenesis-related genes in lesional areas, which attenuates in perilesional skin and is only partially retained in non-lesional areas, validates both sampling accuracy and assay sensitivity. Additionally, perilesional skin displayed prominent upregulation of inflammatory transcripts such as *IFI44* and *IFI6*, consistent with perilesional skin being the zone of highest inflammatory activity in vitiligo (8). Alongside these interferon-inducible genes, we observed strong upregulation of *FOSB*, encoding a transcription factor responsive to stress and inflammatory cues that regulates cell proliferation, differentiation, and transformation (20). A recent study similarly identified *FOSB* as a dominant regulator in vitiligo-associated macrophages, further reinforcing its potential contribution to disease pathogenesis (21).

Across comparisons with healthy controls, lipid metabolism genes were consistently suppressed, supporting the immunometabolic component of vitiligo (22,23). Additional novel findings point to previously unexplored molecular mechanisms: downregulation of *PPP1R1A*, a protein phosphatase-1 (PP1) inhibitor, suggests altered PP1/T-cell receptor signaling, while upregulation of *FAT1* (linked to pro-inflammatory signaling in glioma) and *HORMAD1* (associated with NF-κB activation) further highlight inflammatory networks (24–26). Interestingly, no DEGs were detected between perilesional and non-lesional skin (P_adj_ < 0.05, |log_₂_FC| ≥ 1), likely reflecting the 1 cm sampling proximity and the variable extension of the inflammatory border beyond visible depigmentation, which can blur molecular distinctions (27). These results indicate that nearby non-lesional skin already harbors early pathogenic signals, consistent with prior reports, and suggest that differences observed in more distant regions or compared with healthy controls represent true biological variation rather than technical noise (28).

### Immune and metabolic pathways

Pathway-level analysis highlighted canonical Th1/interferon gamma (IFN-γ)-driven inflammation in vitiligo skin and revealed additional underexplored processes with potential pathogenic relevance. Positive enrichment of NLR and antigen-presentation pathways in both perilesional and non-lesional skin suggests broad activation of immune circuits beyond visibly affected areas. Moreover, enrichment of neutrophil extracellular trap (NET) formation, a process known to drive autoimmunity in psoriasis, indicates a similar pathogenic contribution in vitiligo (29,30).

In parallel, negative enrichment of oxidative phosphorylation, retinol metabolism, and AMP-activated protein kinase (AMPK) signaling points to metabolic dysfunction in vitiligo skin. Since oxidative phosphorylation takes place in mitochondria, this finding is consistent with mitochondrial dysfunction and its correlated increased reactive oxygen species (ROS) production reported in vitiligo, promoting oxidative stress, melanocyte damage, and secondary immune activation (31). Reduced retinol metabolism may reflect a loss of retinoid-mediated protection against oxidative stress and immune activation, while diminished AMPK activity, through loss of autophagy support and mammalian target of rapamycin complex 1 (mTORC1) restraint, potentially exacerbates melanocyte vulnerability (32,33).

### Cellular clearance and homeostasis pathways

Importantly, this study also identifies altered expression of cellular clearance and homeostatic pathways, including ferroptosis, peroxisome, phagosome, and efferocytosis signaling. Impaired clearance of apoptotic or stressed cells may sustain antigen exposure, perpetuating local immune activation and chronicity (34–37). Although these pathways have received little attention in vitiligo, their dysregulation provides a mechanistic link between oxidative stress, defective cell removal, and persistent inflammation. Such findings expand the current immune-centric view of vitiligo to encompass defects in tissue homeostasis and repair.

### Cell growth and structural regulation

Pathways associated with cell growth and structural remodeling were also altered in vitiligo skin. The Hippo signaling pathway, which regulates cell proliferation, differentiation, and survival, was positively enriched in perilesional skin compared to controls (38). Dysregulation of this pathway has been implicated in immune dysfunction and melanocyte dedifferentiation, leading to loss of epidermal identity and impaired repigmentation (39). Its activation at sites of active inflammation in our dataset supports its role in melanocyte dysfunction and resistance to regenerative cues.

The extracellular matrix (ECM)–receptor pathway was positively enriched in lesional skin compared to both perilesional and control skin. It comprises molecules linking the ECM to the cytoskeleton and regulating adhesion, migration, and survival (40,41). Contrary to previous reports of reduced integrin expression in vitiligo, our data showed no such negative enrichment (42). Instead, increased ECM–receptor signaling in lesional skin may reflect inflammatory remodeling, as several of these upregulated pathway genes such as *ITGAV* (log_2_FC = 0.460; P_adj_<0.001), *ITGA11* (log_2_FC = 0.450; P_adj_ > 0.05), *HMMR* (log_2_FC = 0.702; P_adj_ < 0.05), and *COMP* (log_2_FC = 1.778; P_adj_ < 0.001) are typically induced during chronic inflammation and tissue repair (43–45). This remodeling could coexist with local adhesion defects described in previous work, highlighting a more complex regulation of ECM interactions in vitiligo pathogenesis (42).

### Mechanistically distinct candidate compounds point to multiple actionable pathways in vitiligo

The CMap analysis further contextualizes these transcriptional patterns in a therapeutic framework, identifying compounds predicted to reverse the vitiligo-associated signature. Epigenetic modulators emerged as strong candidates, consistent with the role of histone-modification imbalance in melanocyte apoptosis (46). Growth factor kinase inhibitors targeting EGFR, VEGFR, and related non-JAK tyrosine kinases emerged as additional hits, reflecting their cytokine-modulating capacity, yet their use outside oncology remains constrained by dermatologic and vascular toxicities (47–49). Immune signaling modulators targeting BTK, S K, and interleukin pathways further reflect interference with key inflammatory cascades, although compounds such as STA-5326 may also disrupt endolysosomal homeostasis (50,51). Finally, metabolic modulators such as PDE and SphK inhibitors exhibit both anti-inflammatory and pro-melanogenic potential, aligning with early clinical evidence for PDE inhibitors in promoting repigmentation (52–55). Although these results are hypothesis-generating, they illustrate how small-sample transcriptomics can directly inform mechanism-based drug discovery in vitiligo.

## Conclusion

This study demonstrates that 1 mm punch biopsies enable minimally invasive, high-fidelity bulk transcriptomic profiling of vitiligo skin. The resulting data confirm known immune signatures, reveal underexplored structural regulation and clearance pathways, and identify candidate pharmacological modulators, including epigenetic, metabolic and immune-targeted compounds. This small-biopsy transcriptomics approach delivers high-resolution molecular profiling, bridging molecular discovery with clinical application.

## Data Availability

All data produced in the present study are available upon reasonable request to the authors.

## Acknowledgements

The authors express their sincere gratitude to all patients and healthy participants for their valuable participation and contribution to this study. The authors also acknowledge Sylvie Decraene, Sarah De Keulenaer and Ellen De Meester of the Ghent University NXTGNT sequencing core facility for their technical expertise and assistance. We are grateful for their contribution to the Element AVITI sequencing runs, and, in particular, to Sylvie Decraene for performing the Lexogen library preparations.

## References

1. Seneschal J. Clinical Features of Vitiligo and Social Impact on Quality of Life. Dermatol Pract Concept [Internet]. 2023 [cited 2025 May 14];13(4 Suppl 2):e2023312S. Available from: https://pmc.ncbi.nlm.nih.gov/articles/PMC10824319/

2. Speeckaert R, Caelenberg E Van, Belpaire A, Speeckaert MM, Geel N van. Vitiligo: From Pathogenesis to Treatment. J Clin Med [Internet]. 2024 Sep 1 [cited 2025 May 14];13(17). Available from: https://pubmed.ncbi.nlm.nih.gov/39274437/

3. Pathak GN, Tan IJ, Bai G, Dhillon J, Rao BK. Vitiligo: From mechanisms of disease to treatable pathways. Skin Health and Disease [Internet]. 2024 Dec 1 [cited 2025 May 14];4(6). Available from: 10.1002/ski2.460

4. Witek A, Parys J, Mikosińska A, Kaźmierczak M, Mossakowski M, Kałuziak P, et al. Vitiligo Review: etiopathogenesis, diagnosis and treatment. Quality in Sport [Internet]. 2025 Jan 13 [cited 2025 May 14];37:57367. Available from: https://apcz.umk.pl/QS/article/view/57367

5. Seremet T, Di Domizio J, Girardin A, Yatim A, Jenelten R, Messina F, et al. Immune modules to guide diagnosis and personalized treatment of inflammatory skin diseases. Nature Communications 2024 15:1 [Internet]. 2024 Dec 18 [cited 2025 May 21];15(1):1–12. Available from: https://www.nature.com/articles/s41467-024-54559-6

6. Kalmar JR. Advances in the Detection and Diagnosis of Oral Precancerous and Cancerous Lesions. Oral Maxillofac Surg Clin North Am. 2006 Nov;18(4):465–82.

7. Liu D, Hu BD, Mishra A, Patel D, Lau M, Navrazhina K, et al. Tape strips in inflammatory skin disease: a noninvasive method for molecular insights and personalized care. British Journal of Dermatology [Internet]. 2025 Jul 14 [cited 2025 Oct 3];00:1–8. Available from: 10.1093/bjd/ljaf275

8. Martins C, Migayron L, Drullion C, Jacquemin C, Lucchese F, Rambert J, et al. Vitiligo Skin T Cells Are Prone to Produce Type 1 and Type 2 Cytokines to Induce Melanocyte Dysfunction and Epidermal Inflammatory Response Through Jak Signaling. Journal of Investigative Dermatology [Internet]. 2022 Apr 1 [cited 2025 Oct 6];142(4):1194-1205.e7. Available from: https://www.sciencedirect.com/science/article/pii/S0022202X21023046

9. Tulic MK, Kovacs D, Bastonini E, Briganti S, Passeron T, Picardo M. Focusing on the Dark Side of the Moon: Involvement of the Nonlesional Skin in Vitiligo. Journal of Investigative Dermatology [Internet]. 2024 Dec 20 [cited 2025 Jun 16]; Available from: https://www.sciencedirect.com/science/article/pii/S0022202X24028860

10. Brunner PM, David E, Del Duca E, Manson M, Kurowski A, Naidu MP, et al. Transcriptomic profiling of vitiligo patients shows polar immune dysregulation in involved and uninvolved skin. Journal of Allergy and Clinical Immunology [Internet]. 2025 Jun 11 [cited 2025 Jun 16]; Available from: https://linkinghub.elsevier.com/retrieve/pii/S0091674925006268

11. van Geel N, Depaepe L, Vandaele V, Mertens L, Van Causenbroeck J, De Schepper S, et al. Assessing the dynamic changes in vitiligo: reliability and validity of the Vitiligo Disease Activity Score (VDAS) and Vitiligo Disease Improvement Score (VDIS). Journal of the European Academy of Dermatology and Venereology [Internet]. 2022 Aug 1 [cited 2025 Jul 20];36(8):1334–41. Available from: https://pubmed.ncbi.nlm.nih.gov/35398942/

12. Wingett SW, Andrews S. FastQ Screen: A tool for multi-genome mapping and quality control. F1000Res [Internet]. 2018 Sep 17 [cited 2025 Nov 19];7:1338. Available from: https://pubmed.ncbi.nlm.nih.gov/30254741/

13. Martin M. Cutadapt removes adapter sequences from high-throughput sequencing reads. EMBnet J [Internet]. 2011 May 2 [cited 2025 Nov 19];17(1):10–2. Available from: https://journal.embnet.org/index.php/embnetjournal/article/view/200/479

14. Dobin A, Davis CA, Schlesinger F, Drenkow J, Zaleski C, Jha S, et al. STAR: ultrafast universal RNA-seq aligner. Bioinformatics [Internet]. 2013 Jan [cited 2025 Nov 19];29(1):15–21. Available from: https://pubmed.ncbi.nlm.nih.gov/23104886/

15. Smith T, Heger A, Sudbery I. UMI-tools: modeling sequencing errors in Unique Molecular Identifiers to improve quantification accuracy. Genome Res [Internet]. 2017 Mar 1 [cited 2025 Nov 19];27(3):491–9. Available from: https://pubmed.ncbi.nlm.nih.gov/28100584/

16. Li B, Dewey CN. RSEM: accurate transcript quantification from RNA-Seq data with or without a reference genome. BMC Bioinformatics 2011 12:1 [Internet]. 2011 Aug 4 [cited 2025 Nov 19];12(1):323-. Available from: https://bmcbioinformatics.biomedcentral.com/articles/10.1186/1471-2105-12-323

17. Chen Y, Chen L, Lun ATL, Baldoni PL, Smyth GK. edgeR v4: powerful differential analysis of sequencing data with expanded functionality and improved support for small counts and larger datasets. Nucleic Acids Res [Internet]. 2025 Jan 27 [cited 2025 Nov 19];53(2). Available from: https://pubmed.ncbi.nlm.nih.gov/39844453/

18. Yu G, Wang LG, Han Y, He QY. clusterProfiler: an R package for comparing biological themes among gene clusters. OMICS [Internet]. 2012 May 1 [cited 2025 Nov 19];16(5):284–7. Available from: https://pubmed.ncbi.nlm.nih.gov/22455463/

19. Fukushima-Nomura A, Kawasaki H, Yashiro K, Obata S, Tanese K, Ebihara T, et al. An unbiased tissue transcriptome analysis identifies potential markers for skin phenotypes and therapeutic responses in atopic dermatitis. Nature Communications 2025 16:1 [Internet]. 2025 Jun 2 [cited 2025 Jul 1];16(1):1–22. Available from: https://www.nature.com/articles/s41467-025-59340-x

20. FOSB FosB proto-oncogene, AP-1 transcription factor subunit [Homo sapiens (human)] - Gene - NCBI [Internet]. [cited 2025 Sep 5]. Available from: https://www.ncbi.nlm.nih.gov/gene?Db=geneCmd=ShowDetailViewTermToSearch=2354

21. Yu Y, Wang Y, Lu J, Cao X, Feng Y, Pei T, et al. A Comparative Analysis Dissecting the Immune Landscape of Vitiligo and Melanoma from a single-cell Perspective: Two Sides of the Same Coin? Inflammation [Internet]. 2025 Jul 3 [cited 2025 Sep 17];1–19. Available from: https://link.springer.com/article/10.1007/s10753-025-02332-2

22. Glass CK, Olefsky JM. Inflammation and lipid signaling in the etiology of insulin resistance. Cell Metab [Internet]. 2012 May 2 [cited 2025 Sep 17];15(5):635–45. Available from: https://pubmed.ncbi.nlm.nih.gov/22560216/

23. Lyu C, Sun Y. Immunometabolism in the pathogenesis of vitiligo. Front Immunol. 2022 Nov 10;13:1055958.

24. Wu CS, Liu FC, Lin SC, Chyuan IT. Regulation of T cell receptor (TCR) signaling by tyrosine phosphatases: Recent advances and implication for therapeutic approach in autoimmune diseases. Journal of the Formosan Medical Association [Internet]. 2025 Apr 25 [cited 2025 Sep 18]; Available from: https://www.sciencedirect.com/science/article/pii/S0929664625001925

25. Arora M, Kundu A, Sinha S, Chosdol K. Immune factors and their role in tumor aggressiveness in glioblastoma: Atypical cadherin FAT1 as a promising target for combating immune evasion. Cell Mol Biol Lett [Internet]. 2025 Dec 1 [cited 2025 Sep 18];30(1):89. Available from: https://pmc.ncbi.nlm.nih.gov/articles/PMC12291518/

26. Liu K, Cheng L, Zhu K, Wang J, Shu Q. The cancer/testis antigen HORMAD1 mediates epithelial-mesenchymal transition to promote tumor growth and metastasis by activating the Wnt/β-catenin signaling pathway in lung cancer. Cell Death Discov [Internet]. 2022 Mar 28 [cited 2025 Oct 6];8(1):136. Available from: http://www.ncbi.nlm.nih.gov/pubmed/35347116

27. Aslanian FMNP, Noe RAM, Antelo DP, Farias RE, Das PK, Galadari I, et al. Immunohistochemical Findings in Active Vitiligo Including Depigmenting Lesions and Non-Lesional Skin. Open Dermatol J. 2008 Dec 26;2(1):105–10.

28. Tulic MK, Kovacs D, Bastonini E, Briganti S, Passeron T, Picardo M. Focusing on the Dark Side of the Moon: Involvement of the Nonlesional Skin in Vitiligo. Journal of Investigative Dermatology [Internet]. 2024 Jul 1 [cited 2025 Jun 30];145(7). Available from: https://pubmed.ncbi.nlm.nih.gov/39708041/

29. Shao S, Fang H, Dang E, Xue K, Zhang J, Li B, et al. Neutrophil extracellular traps promote inflammatory responses in psoriasis via activating epidermal TLR4/IL-36R crosstalk. Front Immunol [Internet]. 2019 [cited 2025 Sep 29];10(APR):746. Available from: https://pmc.ncbi.nlm.nih.gov/articles/PMC6460719/

30. Wang H, Kim SJ, Lei Y, Wang S, Wang H, Huang H, et al. Neutrophil extracellular traps in homeostasis and disease. Signal Transduct Target Ther [Internet]. 2024 Dec 1 [cited 2025 Sep 29];9(1):1–40. Available from: https://www.nature.com/articles/s41392-024-01933-x

31. Lin Y, Ding Y, Wu Y, Yang Y, Liu Z, Xiang L, et al. The underestimated role of mitochondria in vitiligo: From oxidative stress to inflammation and cell death. Exp Dermatol [Internet]. 2024 Jan 1 [cited 2025 Sep 30];33(1):e14856. Available from: 10.1111/exd.14856

32. Li G, Qu B, Zheng T, Cheng Y, Li P, Liu Z, et al. Assessing the causal effect of genetically predicted metabolites and metabolic pathways on vitiligo: Evidence from Mendelian randomization and animal experiments. J Steroid Biochem Mol Biol [Internet]. 2025 Mar 1 [cited 2025 Sep 30];247:106677. Available from: https://www.sciencedirect.com/science/article/pii/S0960076025000056

33. Bastonini E, Kovacs D, Raffa S, delle Macchie M, Pacifico A, Iacovelli P, et al. A protective role for autophagy in vitiligo. Cell Death Dis [Internet]. 2021 Apr 1 [cited 2025 Sep 30];12(4):1–15. Available from: https://www.nature.com/articles/s41419-021-03592-0

34. Mantegazza AR, Magalhaes JG, Amigorena S, Marks MS. Presentation of phagocytosed antigens by MHC class I and II. Traffic [Internet]. 2012 Feb [cited 2025 Oct 1];14(2):135. Available from: https://pmc.ncbi.nlm.nih.gov/articles/PMC3538944/

35. Tiew TWY, Sheahan MB, Rose RJ. Peroxisomes contribute to reactive oxygen species homeostasis and cell division induction in Arabidopsis protoplasts. Front Plant Sci [Internet]. 2015 Aug 26 [cited 2025 Oct 1];6(AUG):151070. Available from: www.frontiersin.org

36. Meghnem D, Leong E, Pinelli M, Marshall JS, Di Cara F. Peroxisomes Regulate Cellular Free Fatty Acids to Modulate Mast Cell TLR2, TLR4, and IgE-Mediated Activation. Front Cell Dev Biol [Internet]. 2022 May 13 [cited 2025 Oct 1];10:856243. Available from: www.frontiersin.org

37. Li J, Cao F, Yin H liang, Huang Z jian, Lin Z tao, Mao N, et al. Ferroptosis: past, present and future. Cell Death Dis [Internet]. 2020 Feb 1 [cited 2025 Oct 1];11(2):1–13. Available from: https://www.nature.com/articles/s41419-020-2298-2

38. Fu M, Hu Y, Lan T, Guan KL, Luo T, Luo M. The Hippo signalling pathway and its implications in human health and diseases. Signal Transduct Target Ther [Internet]. 2022 Dec 1 [cited 2025 Oct 1];7(1):1–20. Available from: https://www.nature.com/articles/s41392-022-01191-9

39. Yang F, Yang L, Lai S, Yokota M, Kuroda Y, Yuki T, et al. Aberrant laminin signaling drives melanocyte dedifferentiation and unveils a tractable therapeutic target in vitiligo. bioRxiv [Internet]. 2025 Sep 29 [cited 2025 Oct 2];2025.04.11.648350. Available from: https://www.biorxiv.org/content/10.1101/2025.04.11.648350v2

40. KEGG PATHWAY: ECM-receptor interaction - Homo sapiens (human) [Internet]. [cited 2025 Oct 2]. Available from: https://www.kegg.jp/pathway/hsa04512

41. Yue B. Biology of the Extracellular Matrix: An Overview. J Glaucoma [Internet]. 2014 Dec 10 [cited 2025 Oct 2];23(8):S20. Available from: https://pmc.ncbi.nlm.nih.gov/articles/PMC4185430/

42. Su M, Yi H, He X, Luo L, Jiang S, Shi Y. miR-9 regulates melanocytes adhesion and migration during vitiligo repigmentation induced by UVB treatment. Exp Cell Res [Internet]. 2019 Nov 1 [cited 2025 Oct 2];384(1). Available from: https://pubmed.ncbi.nlm.nih.gov/31499059/

43. Schulz JN, Zeltz C, Sørensen IW, Barczyk M, Carracedo S, Hallinger R, et al. Reduced Granulation Tissue and Wound Strength in the Absence of α11β1 Integrin. Journal of Investigative Dermatology [Internet]. 2015 May 1 [cited 2025 Oct 2];135(5):1435–44. Available from: https://www.sciencedirect.com/science/article/pii/S0022202X15372535

44. Jaskuła K, Sacharczuk M, Gaciong Z, Skiba DS. Cardiovascular Effects Mediated by HMMR and CD44. Mediators Inflamm [Internet]. 2021 [cited 2025 Oct 2];2021:4977209. Available from: https://pmc.ncbi.nlm.nih.gov/articles/PMC8286199/

45. Carlsén S, Hansson AS, Olsson H, Heinegård D, Holmdahl R. Cartilage oligomeric matrix protein (COMP)-induced arthritis in rats. Clin Exp Immunol [Internet]. 2001 Dec 25 [cited 2025 Oct 2];114(3):477–84. Available from: 10.1046/j.1365-2249.1998.00739.x

46. Wu L, Han T, Wang Y, Li S, Li C. Epigenetic regulation in vitiligo: mechanisms, challenges, and therapeutic opportunities. Curr Opin Immunol [Internet]. 2025 Aug 1 [cited 2025 Oct 23];95:102580. Available from: https://www.sciencedirect.com/science/article/pii/S0952791525000561

47. Büchau AS. EGFR (Trans)activation Mediates IL-8 and Distinct Human Antimicrobial Peptide and Protein Production following Skin Injury. Journal of Investigative Dermatology [Internet]. 2010 Apr 1 [cited 2025 Oct 23];130(4):929–32. Available from: https://www.sciencedirect.com/science/article/pii/S0022202X15347722

48. Santorsola M, Capuozzo M, Nasti G, Sabbatino F, Di Mauro A, Di Mauro G, et al. Exploring the Spectrum of VEGF Inhibitors’ Toxicities from Systemic to Intra-Vitreal Usage in Medical Practice. Cancers (Basel) [Internet]. 2024 Jan 1 [cited 2025 Oct 23];16(2):350. Available from: https://pmc.ncbi.nlm.nih.gov/articles/PMC10813960/

49. Li T, Perez-Soler R. Skin toxicities associated with epidermal growth factor receptor inhibitors. Target Oncol [Internet]. 2009 Apr [cited 2025 Oct 23];4(2):107–19. Available from: https://pubmed.ncbi.nlm.nih.gov/19452131/

50. Wada Y, Lu R, Zhou D, Chu J, Przewloka T, Zhang S, et al. Selective abrogation of Th1 response by STA-5326, a potent IL-12/IL-23 inhibitor. Blood [Internet]. 2007 Feb 1 [cited 2025 Oct 23];109(3):1156–64. Available from: https://pubmed.ncbi.nlm.nih.gov/17053051/

51. Gayle S, Landrette S, Beeharry N, Conrad C, Hernandez M, Beckett P, et al. Identification of apilimod as a first-in-class PIKfyve kinase inhibitor for treatment of B-cell non-Hodgkin lymphoma. Blood [Internet]. 2017 Mar 30 [cited 2025 Oct 23];129(13):1768–78. Available from: https://www.sciencedirect.com/science/article/pii/S0006497120335369

52. Kim HH, Liao JK. Translational Therapeutics of Dipyridamole. Arterioscler Thromb Vasc Biol [Internet]. 2008 Mar 1 [cited 2025 Oct 23];28(3):s39. Available from: https://pmc.ncbi.nlm.nih.gov/articles/PMC2615560/

53. Chen Z, Li Y, Xie Y, Nie S, Chen B, Wu Z. Roflumilast enhances the melanogenesis and attenuates oxidative stress-triggered damage in melanocytes. J Dermatol Sci [Internet]. 2023 May 1 [cited 2025 Oct 23];110(2):44–52. Available from: https://www.sciencedirect.com/science/article/pii/S0923181123000804

54. Huff SB, Gottwald LD. Repigmentation of Tenacious Vitiligo on Apremilast. Case Rep Dermatol Med. 2017;2017:1–3.

55. Tam I, Kahn JS, Rosmarin D. Repigmentation in a patient with vitiligo on crisaborole 2% ointment. JAAD Case Rep [Internet]. 2021 May 1 [cited 2025 Oct 23];11:99. Available from: https://pmc.ncbi.nlm.nih.gov/articles/PMC8079958/

