## Supplemental Figure S1-S3 for "Minimally invasive 1 mm skin biopsies capture regional transcriptomic heterogeneity in vitiligo"

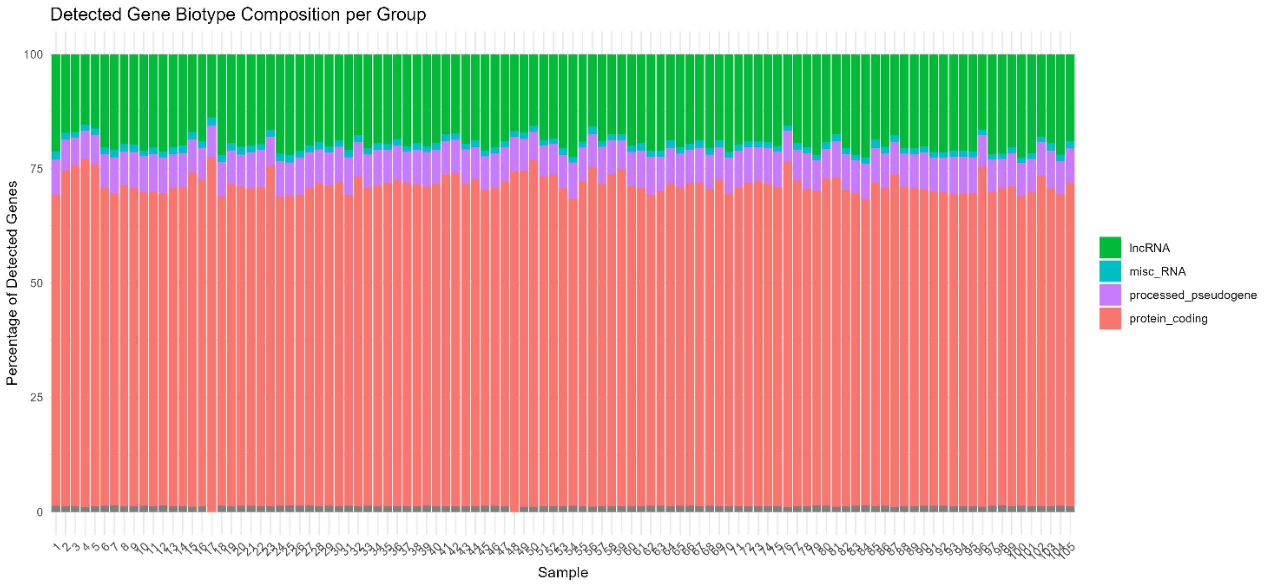


**Figure S1.** Relative composition of detected gene biotypes per sample. Stacked barplots display the proportion of protein-coding, lncRNA, and other noncoding gene types based on the obtained count table. On average, 15,012 protein-coding genes were detected per sample, representing the majority of mapped features.

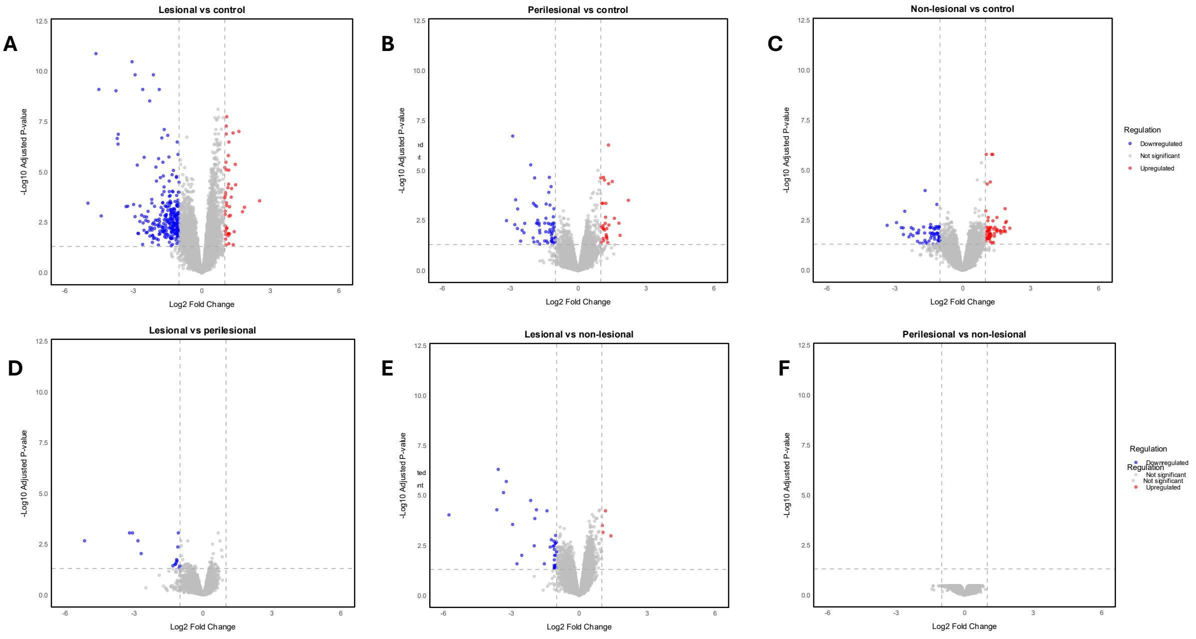


**Figure S2.** Each panel shows a pairwise comparison between skin regions from vitiligo patients and healthy controls: (A) Lesional vs healthy control, (B) Perilesional vs healthy control, (C) Non-lesional vs healthy control, (D) Lesional vs perilesional, (E) Lesional vs non-lesional, (F) Perilesional vs non-lesional. Genes with adjusted p-value < 0.05 and $|\log_2$ fold change$| \geq 1$ are highlighted in red (upregulated) or blue (downregulated), while non-significant genes are shown in grey. Axes represent $\log_2$ fold change (x-axis) and $-\log_{10}$ adjusted p-value (y-axis).

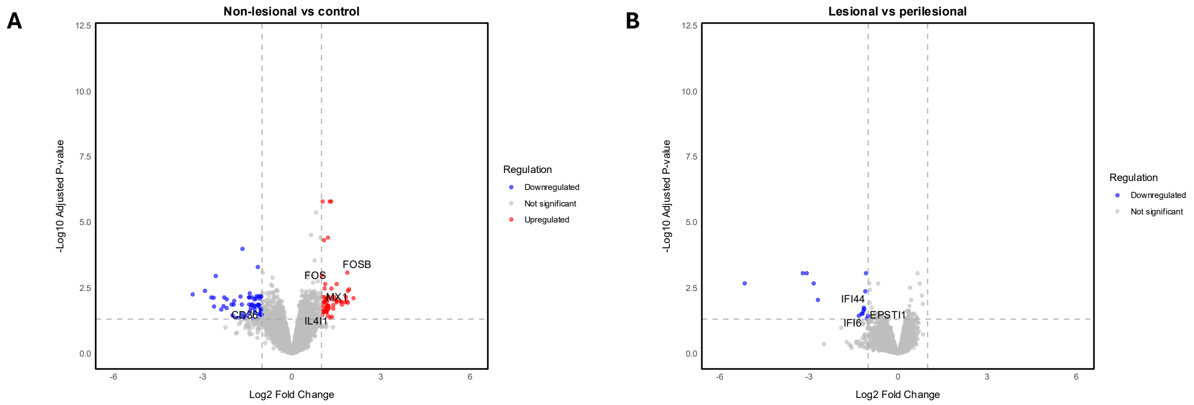


**Figure S3.** Volcano plots highlighting immune-related transcriptional changes in vitiligo skin. Volcano plots display differentially expressed genes in A) non-lesional versus control and B) lesional versus perilesional skin. Immune-related genes including cytokines, chemokines, interferon-stimulated genes, transcription factors, and immune modulators, are annotated where differentially expressed. Blue = downregulated genes, red = upregulated genes, grey = non-significant genes.
