## Supplemental Table S1-S4 for "Minimally invasive 1 mm skin biopsies capture regional transcriptomic heterogeneity in vitiligo"

**Table S1.** Summary of differentially expressed genes (DEGs) identified between patient skin samples and controls. Numbers indicate total DEGs as well as genes up- or down-regulated in group A relative to group B.

| group A (# repl.) | group B (# repl.) | differential A vs B | up-regulated A | down-regulated A |
|---|---|---|---|---|
| Lesional (24) | Control (27) | 320 | 45 | 275 |
| Perilesional (27) | Control (27) | 92 | 30 | 62 |
| Non-lesional (27) | Control (27) | 124 | 63 | 61 |
| Lesional (24) | Perilesional (27) | 15 | 0 | 15 |
| Lesional (24) | Non-lesional (27) | 40 | 4 | 36 |
| Perilesional (27) | Non-lesional (27) | 0 | 0 | 0 |

**Table S2.** Gene Ontology (GO) enrichment results for biological processes (BP) in the lesional versus control comparison, filtered for pathways with normalized enrichment score (NES) ≥ 2 and adjusted p < 0.05. GO:0045814; negative regulation of gene expression, epigenetic; is highlighted in bold.

| ID | Description | NES | $P_{adj}$ |
|---|---|---|---|
| GO:0007062 | sister chromatid cohesion | 2,4080764 | 1,1491E-05 |
| GO:1905818 | regulation of chromosome separation | 2,37586239 | 5,8777E-06 |
| GO:0051983 | regulation of chromosome segregation | 2,33556994 | 1,2129E-07 |
| GO:0051304 | chromosome separation | 2,32029497 | 7,2355E-06 |
| GO:0051784 | negative regulation of nuclear division | 2,30408687 | 3,5148E-05 |
| GO:0010965 | regulation of mitotic sister chromatid separation | 2,2584731 | 9,4035E-05 |
| GO:0051321 | meiotic cell cycle | 2,25599636 | 1,0352E-08 |
| GO:0070192 | chromosome organization involved in meiotic cell cycle | 2,21415731 | 0,00047958 |
| GO:0045839 | negative regulation of mitotic nuclear division | 2,21210956 | 0,00030294 |
| GO:0033046 | negative regulation of sister chromatid segregation | 2,21068349 | 0,00054757 |
| GO:0033048 | negative regulation of mitotic sister chromatid segregation | 2,21068349 | 0,00054757 |
| GO:0045841 | negative regulation of mitotic metaphase/anaphase transition | 2,21068349 | 0,00054757 |
| GO:2000816 | negative regulation of mitotic sister chromatid separation | 2,21068349 | 0,00054757 |
| GO:0007094 | mitotic spindle assembly checkpoint signaling | 2,20471542 | 0,00070023 |
| GO:0071173 | spindle assembly checkpoint signaling | 2,20471542 | 0,00070023 |
| GO:0071174 | mitotic spindle checkpoint signaling | 2,20471542 | 0,00070023 |
| GO:0051985 | negative regulation of chromosome segregation | 2,1801941 | 0,00054597 |
| GO:1902100 | negative regulation of metaphase/anaphase transition of cell cycle | 2,1801941 | 0,00054597 |
| GO:1905819 | negative regulation of chromosome separation | 2,1801941 | 0,00054597 |
| GO:0031577 | spindle checkpoint signaling | 2,17769444 | 0,00070023 |
| GO:0007059 | chromosome segregation | 2,17391395 | 3,809E-14 |
| GO:0051383 | kinetochore organization | 2,16937553 | 0,00469969 |
| GO:0009649 | entrainment of circadian clock | 2,1677217 | 0,00265801 |
| GO:0007064 | mitotic sister chromatid cohesion | 2,16216502 | 0,00104778 |
| GO:0009648 | photoperiodism | 2,16008956 | 0,00372215 |
| GO:0043153 | entrainment of circadian clock by photoperiod | 2,15903808 | 0,00524317 |
| GO:0071459 | protein localization to chromosome, centromeric region | 2,15708684 | 0,00224621 |
| **GO:0045814** | **negative regulation of gene expression, epigenetic** | **2,14721573** | **5,6066E-06** |
| GO:0031507 | heterochromatin formation | 2,14696455 | 7,704E-05 |
| GO:0033047 | regulation of mitotic sister chromatid segregation | 2,14585394 | 0,00058593 |
| GO:0051783 | regulation of nuclear division | 2,12809564 | 5,8725E-06 |
| GO:0000070 | mitotic sister chromatid segregation | 2,12100351 | 1,9979E-07 |
| GO:0098813 | nuclear chromosome segregation | 2,11784954 | 9,3075E-10 |
| GO:0045132 | meiotic chromosome segregation | 2,11315381 | 0,00065023 |
| GO:0051306 | mitotic sister chromatid separation | 2,10233024 | 0,00036055 |
| GO:0070828 | heterochromatin organization | 2,09633721 | 0,00022772 |
| GO:0090068 | positive regulation of cell cycle process | 2,09301778 | 3,9915E-08 |
| GO:1903046 | meiotic cell cycle process | 2,08767765 | 1,0607E-05 |
| GO:0034508 | centromere complex assembly | 2,08431039 | 0,00519814 |

| | | | |
|---|---|---|---|
| GO:1902969 | mitotic DNA replication | 2,05302471 | 0,00640999 |
| GO:0000280 | nuclear division | 2,04474979 | 6,8794E-11 |
| GO:0051642 | centrosome localization | 2,04329187 | 0,00623354 |
| GO:0061842 | microtubule organizing center localization | 2,04329187 | 0,00623354 |
| GO:0007088 | regulation of mitotic nuclear division | 2,03279994 | 0,00011483 |
| GO:0018149 | peptide cross-linking | 2,0227442 | 0,01125199 |

**Table S3:** Ranked list of KEGG pathways for all comparisons, ordered by adjusted p-value ($P_{adj}$). For each pathway, $P_{adj}$, normalized enrichment score (NES), and specific comparison are shown. The pathways discussed in the main text of the manuscript are highlighted in bold. L, lesional skin; P, perilesional skin; NL, non-lesional skin; C, control skin.

| KEGG pathway | Comparison | NES | $P_{adj}$ |
|---|---|---|---|
| Diabetic cardiomyopathy | L vs C | -2,41582573 | 3,8226E-15 |
| **PPAR signaling pathway** | **L vs C** | **-2,66466697** | **6,2221E-14** |
| **Oxidative phosphorylation** | **L vs C** | **-2,41428574** | **2,8919E-12** |
| Thermogenesis | L vs C | -2,20058958 | 4,0836E-12 |
| Non-alcoholic fatty liver disease | L vs C | -2,1863408 | 4,2559E-09 |
| Chemical carcinogenesis - reactive oxygen species | L vs C | -2,09826115 | 4,2559E-09 |
| **Peroxisome** | **L vs C** | **-2,29556625** | **5,921E-08** |
| Parkinson disease | L vs C | -1,93398027 | 2,5522E-07 |
| **Fatty acid metabolism** | **L vs C** | **-2,26833261** | **1,8563E-06** |
| Cardiac muscle contraction | L vs C | -2,24603098 | 1,9033E-06 |
| Prion disease | L vs C | -1,88553467 | 3,1856E-06 |
| **Cell cycle** | **L vs C** | **2,08123762** | **7,072E-06** |
| Huntington disease | L vs C | -1,7896498 | 1,2774E-05 |
| Fatty acid degradation | L vs C | -2,18029752 | 1,8198E-05 |
| Valine, leucine and isoleucine degradation | L vs C | -2,15421245 | 2,9439E-05 |
| **Tyrosine metabolism** | **L vs C** | **-2,16892** | **3,2613E-05** |
| Carbon metabolism | L vs C | -2,0297161 | 3,4368E-05 |
| Spliceosome | L vs C | 1,93385383 | 6,4743E-05 |
| Alzheimer disease | L vs C | -1,65159445 | 6,4743E-05 |
| Biosynthesis of unsaturated fatty acids | L vs C | -2,13052202 | 6,4845E-05 |
| Cholesterol metabolism | L vs C | -2,11431675 | 9,1219E-05 |
| 2-Oxocarboxylic acid metabolism | L vs C | -2,14309052 | 9,5531E-05 |
| Glycerolipid metabolism | L vs C | -2,10451837 | 0,00013973 |
| Drug metabolism - cytochrome P450 | L vs C | -2,10684853 | 0,00020421 |
| Pathways of neurodegeneration - multiple diseases | L vs C | -1,56069234 | 0,00037183 |
| Ribosome biogenesis in eukaryotes | L vs C | 1,98128496 | 0,00041918 |
| Circadian rhythm | L vs C | 2,15594034 | 0,00089243 |
| Fatty acid elongation | L vs C | -2,01254123 | 0,00089243 |
| Terpenoid backbone biosynthesis | L vs C | -1,98957779 | 0,00089243 |
| **Adipocytokine signaling pathway** | **L vs C** | **-1,94549279** | **0,00089243** |
| Retrograde endocannabinoid signaling | L vs C | -1,85288274 | 0,00089243 |
| Pyruvate metabolism | L vs C | -1,95842378 | 0,0010105 |
| Steroid biosynthesis | L vs C | -1,95755089 | 0,0010105 |

| | | | |
|---|---|---|---|
| Alcoholic liver disease | L vs C | -1,80110984 | 0,00103502 |
| Malaria | L vs C | -1,95372666 | 0,00126694 |
| Metabolism of xenobiotics by cytochrome P450 | L vs C | -1,97341956 | 0,00132164 |
| Glycolysis / Gluconeogenesis | L vs C | -1,95918928 | 0,00132164 |
| Nucleocytoplasmic transport | L vs C | 1,85517379 | 0,00132164 |
| Insulin resistance | L vs C | -1,83885362 | 0,00132164 |
| Rheumatoid arthritis | L vs C | -1,85923784 | 0,001577 |
| Staphylococcus aureus infection | L vs C | -1,94892266 | 0,00177193 |
| Tryptophan metabolism | L vs C | -1,96601515 | 0,00209692 |
| Citrate cycle (TCA cycle) | L vs C | -1,96464557 | 0,00209692 |
| Intestinal immune network for IgA production | L vs C | -1,96205092 | 0,00209692 |
| **AMPK signaling pathway** | L vs C | -1,77511739 | 0,00246134 |
| Biosynthesis of amino acids | L vs C | -1,91090906 | 0,00275946 |
| African trypanosomiasis | L vs C | -1,88191982 | 0,00292216 |
| Butanoate metabolism | L vs C | -1,89973105 | 0,00305877 |
| Amyotrophic lateral sclerosis | L vs C | -1,50467426 | 0,00380378 |
| Lysosome | L vs C | -1,65660009 | 0,00525824 |
| **Ferroptosis** | **L vs C** | **-1,82177259** | **0,00553054** |
| Steroid hormone biosynthesis | L vs C | -1,85383621 | 0,00634698 |
| **Melanogenesis** | **L vs C** | **-1,73289425** | **0,00825058** |
| Hematopoietic cell lineage | L vs C | -1,79064562 | 0,00908589 |
| Drug metabolism - other enzymes | L vs C | -1,70708857 | 0,01011528 |
| Chemical carcinogenesis - DNA adducts | L vs C | -1,79950187 | 0,01165355 |
| Glycerophospholipid metabolism | L vs C | -1,66781492 | 0,0120273 |
| Glyoxylate and dicarboxylate metabolism | L vs C | -1,85948606 | 0,01203407 |
| Regulation of lipolysis in adipocytes | L vs C | -1,78251802 | 0,01412959 |
| Fatty acid biosynthesis | L vs C | -1,82432898 | 0,01460376 |
| Pentose and glucuronate interconversions | L vs C | -1,78739836 | 0,01483419 |
| Glutathione metabolism | L vs C | -1,76109083 | 0,01483419 |
| Ascorbate and aldarate metabolism | L vs C | -1,73510491 | 0,01486795 |
| Asthma | L vs C | -1,80169183 | 0,01819459 |
| Starch and sucrose metabolism | L vs C | -1,71342098 | 0,0190766 |
| Fat digestion and absorption | L vs C | -1,72110009 | 0,02154922 |
| Phenylalanine metabolism | L vs C | -1,72302918 | 0,02407835 |
| Basal transcription factors | L vs C | 1,70835911 | 0,02492046 |
| Propanoate metabolism | L vs C | -1,69391091 | 0,02752453 |
| Cell adhesion molecules | L vs C | -1,56581012 | 0,03355086 |
| **HIF-1 signaling pathway** | **L vs C** | **-1,59060072** | **0,03394392** |
| Porphyrin metabolism | L vs C | -1,67732974 | 0,03801749 |
| ECM-receptor interaction | L vs C | 1,54493036 | 0,03801749 |
| **Retinol metabolism** | L vs C | -1,61191306 | 0,04802106 |
| Tuberculosis | L vs C | -1,50905412 | 0,04802106 |
| **PPAR signaling pathway** | **P vs C** | **-2,675644928** | **3,82E-12** |
| Diabetic cardiomyopathy | P vs C | -2,369640955 | 1,64E-10 |
| Drug metabolism - cytochrome P450 | P vs C | -2,565279151 | 1,22E-09 |
| **Oxidative phosphorylation** | **P vs C** | **-2,345148973** | **2,18E-08** |

| | | | |
|---|---|---|---|
| **Retinol metabolism** | P vs C | -2,426893732 | 6,21E-08 |
| Carbon metabolism | P vs C | -2,322706107 | 1,62E-07 |
| Thermogenesis | P vs C | -2,120958716 | 1,81E-07 |
| Metabolism of xenobiotics by cytochrome P450 | P vs C | -2,390415505 | 1,82E-07 |
| Pentose and glucuronate interconversions | P vs C | -2,377796631 | 2,34E-07 |
| Non-alcoholic fatty liver disease | P vs C | -2,131831708 | 1,51E-06 |
| Ascorbate and aldarate metabolism | P vs C | -2,278258434 | 1,63E-06 |
| Chemical carcinogenesis - reactive oxygen species | P vs C | -2,033615779 | 1,71E-06 |
| **Tyrosine metabolism** | **P vs C** | **-2,286227451** | **3,48E-06** |
| **Peroxisome** | **P vs C** | **-2,248063796** | **9E-06** |
| Measles | P vs C | 2,073674148 | 1,43E-05 |
| Steroid hormone biosynthesis | P vs C | -2,230637755 | 2,44E-05 |
| Fatty acid metabolism | P vs C | -2,234381128 | 2,79E-05 |
| Cardiac muscle contraction | P vs C | -2,215064524 | 2,79E-05 |
| Glycolysis / Gluconeogenesis | P vs C | -2,246298862 | 4,65E-05 |
| Drug metabolism - other enzymes | P vs C | -2,100594527 | 0,000126 |
| Glycerolipid metabolism | P vs C | -2,180502301 | 0,000156 |
| Spliceosome | P vs C | 1,845480837 | 0,000179 |
| Fatty acid degradation | P vs C | -2,177573927 | 0,000182 |
| Herpes simplex virus 1 infection | P vs C | 1,840505061 | 0,000182 |
| Circadian rhythm | P vs C | 2,12170913 | 0,000256 |
| 2-Oxocarboxylic acid metabolism | P vs C | -2,11549947 | 0,00028 |
| Chemical carcinogenesis - DNA adducts | P vs C | -2,078086968 | 0,00028 |
| Valine, leucine and isoleucine degradation | P vs C | -2,070983091 | 0,00028 |
| **Adipocytokine signaling pathway** | **P vs C** | **-2,063008968** | **0,00028** |
| Parkinson disease | P vs C | -1,783863531 | 0,000306 |
| Porphyrin metabolism | P vs C | -2,052954895 | 0,000314 |
| Pyruvate metabolism | P vs C | -2,011028568 | 0,000381 |
| Biosynthesis of cofactors | P vs C | -1,878918203 | 0,000445 |
| Influenza A | P vs C | 1,812857264 | 0,00049 |
| **AMPK signaling pathway** | P vs C | -1,873140276 | 0,000861 |
| Alzheimer disease | P vs C | -1,624394491 | 0,000882 |
| Leishmaniasis | P vs C | 1,893129321 | 0,000887 |
| Cholesterol metabolism | P vs C | -2,000916519 | 0,000927 |
| Propanoate metabolism | P vs C | -1,986727513 | 0,00186 |
| Citrate cycle (TCA cycle) | P vs C | -1,985271105 | 0,002463 |
| Biosynthesis of amino acids | P vs C | -1,938101379 | 0,002613 |
| Epstein-Barr virus infection | P vs C | 1,654734312 | 0,002944 |
| Bile secretion | P vs C | -1,908590906 | 0,003139 |
| **NOD-like receptor signaling pathway** | **P vs C** | **1,729842038** | **0,003139** |
| Glyoxylate and dicarboxylate metabolism | P vs C | -1,952556135 | 0,003316 |
| Huntington disease | P vs C | -1,642650726 | 0,003316 |
| Phenylalanine metabolism | P vs C | -1,874326606 | 0,003379 |
| Biosynthesis of unsaturated fatty acids | P vs C | -1,960566145 | 0,003593 |

| | | | |
|---|---|---|---|
| Mineral absorption | P vs C | -1,870241698 | 0,003671 |
| Hepatitis C | P vs C | 1,646393345 | 0,00375 |
| **TNF signaling pathway** | **P vs C** | **1,688426335** | **0,00381** |
| Cornified envelope formation | P vs C | 1,630899152 | 0,00381 |
| Regulation of lipolysis in adipocytes | P vs C | -1,874910223 | 0,004087 |
| **IL-17 signaling pathway** | **P vs C** | **1,806377844** | **0,005454** |
| Pentose phosphate pathway | P vs C | -1,890002853 | 0,005557 |
| Alcoholic liver disease | P vs C | -1,767353342 | 0,005557 |
| Steroid biosynthesis | P vs C | -1,885762538 | 0,006056 |
| **Antigen processing and presentation** | **P vs C** | **1,788144238** | **0,007445** |
| Glycerophospholipid metabolism | P vs C | -1,701044151 | 0,007911 |
| Starch and sucrose metabolism | P vs C | -1,783041166 | 0,009332 |
| Central carbon metabolism in cancer | P vs C | -1,771189828 | 0,009332 |
| Pathways of neurodegeneration - multiple diseases | P vs C | -1,44475062 | 0,009398 |
| Ribosome biogenesis in eukaryotes | P vs C | 1,668935471 | 0,01042 |
| Prion disease | P vs C | -1,561794488 | 0,010949 |
| Fatty acid elongation | P vs C | -1,81043411 | 0,011915 |
| Inflammatory bowel disease | P vs C | 1,856533124 | 0,012449 |
| **Melanogenesis** | **P vs C** | **-1,718399295** | **0,01251** |
| Fat digestion and absorption | P vs C | -1,81209772 | 0,013002 |
| Fatty acid biosynthesis | P vs C | -1,809307797 | 0,014733 |
| beta-Alanine metabolism | P vs C | -1,828107338 | 0,015283 |
| Glycine, serine and threonine metabolism | P vs C | -1,846448139 | 0,016139 |
| alpha-Linolenic acid metabolism | P vs C | -1,820362944 | 0,016139 |
| Fructose and mannose metabolism | P vs C | -1,71813263 | 0,022418 |
| Dilated cardiomyopathy | P vs C | -1,678216143 | 0,02441 |
| **HIF-1 signaling pathway** | **P vs C** | **-1,629967832** | **0,02441** |
| Insulin resistance | P vs C | -1,58344309 | 0,02441 |
| Lysosome | P vs C | -1,515198924 | 0,024693 |
| Butanoate metabolism | P vs C | -1,7431382 | 0,026651 |
| **Hippo signaling pathway** | **P vs C** | **1,48194668** | **0,026651** |
| Coronavirus disease - COVID-19 | P vs C | 1,395232305 | 0,029014 |
| **Ferroptosis** | **P vs C** | **-1,638325511** | **0,032869** |
| Galactose metabolism | P vs C | -1,649170141 | 0,033023 |
| Cytoskeleton in muscle cells | P vs C | -1,481014358 | 0,035585 |
| **Cell cycle** | **P vs C** | **1,421646428** | **0,037394** |
| Hypertrophic cardiomyopathy | P vs C | -1,592806717 | 0,037421 |
| **Th1 and Th2 cell differentiation** | **P vs C** | **1,516573973** | **0,043595** |
| Calcium signaling pathway | P vs C | -1,478863665 | 0,045308 |
| **PPAR signaling pathway** | **NL vs C** | **-2,85429602** | **4,98361E-16** |
| **Oxidative phosphorylation** | **NL vs C** | **-2,72932619** | **4,98361E-16** |
| Diabetic cardiomyopathy | NL vs C | -2,56706859 | 7,47543E-16 |
| Chemical carcinogenesis - reactive oxygen species | NL vs C | -2,54054566 | 9,66089E-16 |
| Thermogenesis | NL vs C | -2,48197716 | 5,3541E-15 |
| Carbon metabolism | NL vs C | -2,59431595 | 2,37272E-12 |

| | | | |
|---|---|---|---|
| Non-alcoholic fatty liver disease | NL vs C | -2,48309227 | 3,69876E-12 |
| Parkinson disease | NL vs C | -2,23880605 | 5,87827E-11 |
| **Fatty acid metabolism** | **NL vs C** | **-2,56205636** | **2,57565E-10** |
| **Peroxisome** | **NL vs C** | **-2,49489913** | **1,10199E-09** |
| Huntington disease | NL vs C | -2,06355594 | 2,20285E-08 |
| Citrate cycle (TCA cycle) | NL vs C | -2,48809386 | 3,66421E-08 |
| Prion disease | NL vs C | -2,02600384 | 2,44945E-07 |
| Alzheimer disease | NL vs C | -1,92180741 | 2,44945E-07 |
| Metabolism of xenobiotics by cytochrome P450 | NL vs C | -2,3468712 | 8,49823E-07 |
| Pyruvate metabolism | NL vs C | -2,35869735 | 1,25384E-06 |
| Drug metabolism - cytochrome P450 | NL vs C | -2,35405398 | 1,42056E-06 |
| Amyotrophic lateral sclerosis | NL vs C | -1,87951034 | 2,18936E-06 |
| Glycolysis / Gluconeogenesis | NL vs C | -2,28830545 | 2,36942E-06 |
| 2-Oxocarboxylic acid metabolism | NL vs C | -2,29405602 | 3,53528E-06 |
| Fatty acid degradation | NL vs C | -2,31570325 | 6,93978E-06 |
| Fatty acid elongation | NL vs C | -2,28559426 | 1,80371E-05 |
| Glutathione metabolism | NL vs C | -2,17851672 | 2,09885E-05 |
| Spliceosome | NL vs C | 2,00866821 | 2,15868E-05 |
| Glycerolipid metabolism | NL vs C | -2,18243064 | 2,63909E-05 |
| Biosynthesis of unsaturated fatty acids | NL vs C | -2,23413256 | 3,03393E-05 |
| Chemical carcinogenesis - DNA adducts | NL vs C | -2,2164699 | 3,46514E-05 |
| Glyoxylate and dicarboxylate metabolism | NL vs C | -2,20504001 | 3,46514E-05 |
| Measles | NL vs C | 2,07417705 | 3,52772E-05 |
| Biosynthesis of amino acids | NL vs C | -2,19061393 | 4,15613E-05 |
| **Adipocytokine signaling pathway** | **NL vs C** | **-2,18136849** | **4,92928E-05** |
| Terpenoid backbone biosynthesis | NL vs C | -2,20346064 | 5,4594E-05 |
| Valine, leucine and isoleucine degradation | NL vs C | -2,17034932 | 5,55352E-05 |
| Steroid hormone biosynthesis | NL vs C | -2,15370059 | 7,94654E-05 |
| Propanoate metabolism | NL vs C | -2,16956133 | 0,000102093 |
| Regulation of lipolysis in adipocytes | NL vs C | -2,09644256 | 0,000119251 |
| Motor proteins | NL vs C | 1,93879838 | 0,000133577 |
| **NOD-like receptor signaling pathway** | **NL vs C** | **1,89594667** | **0,0001552** |
| Steroid biosynthesis | NL vs C | -2,11747469 | 0,000191623 |
| Pathways of neurodegeneration - multiple diseases | NL vs C | -1,63765903 | 0,000298373 |
| Butanoate metabolism | NL vs C | -2,11052502 | 0,000382605 |
| **AMPK signaling pathway** | NL vs C | -1,86168619 | 0,000551309 |
| **Retinol metabolism** | NL vs C | -2,02934706 | 0,000741015 |
| Pentose and glucuronate interconversions | NL vs C | -2,09071844 | 0,000890948 |
| Cardiac muscle contraction | NL vs C | -1,97698736 | 0,001113728 |
| **Tyrosine metabolism** | **NL vs C** | **-1,96958082** | **0,001189942** |
| Drug metabolism - other enzymes | NL vs C | -1,87145928 | 0,001782292 |
| Biosynthesis of cofactors | NL vs C | -1,72902726 | 0,001782292 |
| Pentose phosphate pathway | NL vs C | -1,94103696 | 0,001804438 |
| Herpes simplex virus 1 infection | NL vs C | 1,69140554 | 0,001943793 |
| Fatty acid biosynthesis | NL vs C | -1,97843126 | 0,001962484 |
| Ribosome | NL vs C | -1,69049089 | 0,003025813 |

| | | | |
|---|---|---|---|
| Cholesterol metabolism | NL vs C | -1,91931875 | 0,003066107 |
| Influenza A | NL vs C | 1,70163255 | 0,00397229 |
| Epstein-Barr virus infection | NL vs C | 1,59267751 | 0,005032523 |
| Retrograde endocannabinoid signaling | NL vs C | -1,70747947 | 0,005370705 |
| Ascorbate and aldarate metabolism | NL vs C | -1,93547544 | 0,00571823 |
| Fat digestion and absorption | NL vs C | -1,89610072 | 0,005778465 |
| Glycerophospholipid metabolism | NL vs C | -1,72616937 | 0,005778465 |
| Circadian entrainment | NL vs C | 1,81087914 | 0,006035822 |
| Cornified envelope formation | NL vs C | 1,61896825 | 0,006203661 |
| Glycine, serine and threonine metabolism | NL vs C | -1,91061373 | 0,007590132 |
| Mineral absorption | NL vs C | -1,81972351 | 0,011050818 |
| Taste transduction | NL vs C | 1,88717959 | 0,011196306 |
| Alcoholic liver disease | NL vs C | -1,6303599 | 0,013107577 |
| Porphyrin metabolism | NL vs C | -1,81464898 | 0,01408299 |
| **Th1 and Th2 cell differentiation** | **NL vs C** | **1,73230936** | **0,01408299** |
| **Ferroptosis** | **NL vs C** | **-1,76344583** | **0,016864259** |
| Endocytosis | NL vs C | 1,44194651 | 0,017581504 |
| **Antigen processing and presentation** | **NL vs C** | **1,69051557** | **0,019207347** |
| alpha-Linolenic acid metabolism | NL vs C | -1,77451733 | 0,025153056 |
| Lipoic acid metabolism | NL vs C | -1,74522259 | 0,025153056 |
| **TNF signaling pathway** | **NL vs C** | **1,56107992** | **0,025263388** |
| Insulin resistance | NL vs C | -1,56192089 | 0,02586737 |
| Central carbon metabolism in cancer | NL vs C | -1,63793013 | 0,026983217 |
| Systemic lupus erythematosus | NL vs C | 1,64172656 | 0,031219442 |
| Human papillomavirus infection | NL vs C | 1,38465234 | 0,031219442 |
| Human immunodeficiency virus 1 infection | NL vs C | 1,41540125 | 0,037633172 |
| Focal adhesion | NL vs C | 1,40797974 | 0,040082477 |
| beta-Alanine metabolism | NL vs C | -1,73901872 | 0,041378579 |
| Starch and sucrose metabolism | NL vs C | -1,71219305 | 0,04159669 |
| Circadian rhythm | NL vs C | 1,6702569 | 0,042194076 |
| Diabetic cardiomyopathy | L vs P | 2,08982399 | 7,1896E-09 |
| **Oxidative phosphorylation** | **L vs P** | **2,16528715** | **1,1242E-07** |
| Thermogenesis | L vs P | 1,95798619 | 1,1242E-07 |
| Chemical carcinogenesis - reactive oxygen species | L vs P | 1,92284766 | 5,3199E-07 |
| Staphylococcus aureus infection | L vs P | 2,31115379 | 1,3781E-06 |
| **PPAR signaling pathway** | **L vs P** | **2,23407205** | **1,8916E-06** |
| Non-alcoholic fatty liver disease | L vs P | 1,98619171 | 4,9951E-06 |
| Intestinal immune network for IgA production | L vs P | 2,21319497 | 9,6763E-06 |
| Prion disease | L vs P | 1,81807536 | 1,0081E-05 |
| Cell adhesion molecules | L vs P | 2,05072751 | 1,436E-05 |
| Hematopoietic cell lineage | L vs P | 2,10717186 | 6,5977E-05 |
| Huntington disease | L vs P | 1,71493761 | 6,5977E-05 |
| **Fatty acid metabolism** | **L vs P** | **2,13529592** | **6,9548E-05** |
| **Phagosome** | **L vs P** | **1,92186254** | **8,4375E-05** |
| Rheumatoid arthritis | L vs P | 2,09638263 | 8,6659E-05 |
| Ascorbate and aldarate metabolism | L vs P | -2,34875809 | 0,00012188 |

| | | | |
|---|---|---|---|
| Parkinson disease | L vs P | 1,68873393 | 0,00012188 |
| Leishmaniasis | L vs P | 2,03244583 | 0,00035844 |
| Biosynthesis of unsaturated fatty acids | L vs P | 2,07349707 | 0,00044119 |
| Epstein-Barr virus infection | L vs P | 1,76180985 | 0,00061101 |
| Systemic lupus erythematosus | L vs P | 1,9494855 | 0,00085822 |
| Retrograde endocannabinoid signaling | L vs P | 1,85714331 | 0,00085822 |
| Alzheimer disease | L vs P | 1,53073935 | 0,00085822 |
| Graft-versus-host disease | L vs P | 2,00098221 | 0,00093919 |
| Autoimmune thyroid disease | L vs P | 1,99771875 | 0,00093919 |
| Allograft rejection | L vs P | 1,99771875 | 0,00093919 |
| Osteoclast differentiation | L vs P | 1,8329391 | 0,00093919 |
| Influenza A | L vs P | 1,72434048 | 0,00093919 |
| Type I diabetes mellitus | L vs P | 2,00941648 | 0,00102165 |
| **Tyrosine metabolism** | **L vs P** | **1,99080412** | **0,00102165** |
| **Chemokine signaling pathway** | **L vs P** | **1,78922256** | **0,00102165** |
| Tuberculosis | L vs P | 1,74242141 | 0,00112662 |
| **Cell cycle** | **L vs P** | **-1,62881759** | **0,00128914** |
| Complement and coagulation cascades | L vs P | 1,94037714 | 0,00198006 |
| Nucleocytoplasmic transport | L vs P | -1,73229927 | 0,00198006 |
| **Peroxisome** | **L vs P** | **1,83232845** | **0,00209481** |
| Asthma | L vs P | 1,95685639 | 0,00210676 |
| Viral myocarditis | L vs P | 1,86841182 | 0,00226818 |
| Malaria | L vs P | 1,93658821 | 0,00233702 |
| Fatty acid degradation | L vs P | 1,8926906 | 0,00233702 |
| Lysosome | L vs P | 1,72949966 | 0,00233702 |
| Herpes simplex virus 1 infection | L vs P | 1,68427246 | 0,00233702 |
| African trypanosomiasis | L vs P | 1,92381352 | 0,00257005 |
| Pentose and glucuronate interconversions | L vs P | -2,00973055 | 0,00277075 |
| **Efferocytosis** | **L vs P** | **1,71945142** | **0,00279934** |
| Pathways of neurodegeneration - multiple diseases | L vs P | 1,44542146 | 0,00358572 |
| Inflammatory bowel disease | L vs P | 1,80423713 | 0,00512211 |
| **Natural killer cell mediated cytotoxicity** | **L vs P** | **1,79726114** | **0,00692851** |
| **Cytokine-cytokine receptor interaction** | **L vs P** | **1,69587372** | **0,00692851** |
| **AMPK signaling pathway** | **L vs P** | **1,64892678** | **0,00718463** |
| **Antigen processing and presentation** | **L vs P** | **1,80586927** | **0,0076764** |
| **Melanogenesis** | **L vs P** | **1,75708141** | **0,00768496** |
| Terpenoid backbone biosynthesis | L vs P | 1,79622123 | 0,0077341 |
| Cardiac muscle contraction | L vs P | 1,76936725 | 0,0077341 |
| **Viral protein interaction with cytokine and cytokine receptor** | **L vs P** | **1,78350427** | **0,00875462** |
| Salivary secretion | L vs P | 1,78050606 | 0,01018853 |
| Human T-cell leukemia virus 1 infection | L vs P | 1,49323122 | 0,01131755 |
| Motor proteins | L vs P | -1,49449142 | 0,01347539 |
| **Neutrophil extracellular trap formation** | **L vs P** | **1,61683346** | **0,01436647** |
| Chagas disease | L vs P | 1,65440548 | 0,01524752 |
| Drug metabolism - cytochrome P450 | L vs P | -1,67205351 | 0,01662163 |
| Pertussis | L vs P | 1,6836129 | 0,01689983 |

| | | | |
|---|---|---|---|
| **ECM-receptor interaction** | **L vs P** | **-1,66017901** | **0,01689983** |
| Measles | L vs P | 1,64046055 | 0,01703191 |
| Alcoholic liver disease | L vs P | 1,60404809 | 0,01703191 |
| Vibrio cholerae infection | L vs P | 1,66463965 | 0,01807169 |
| Homologous recombination | L vs P | -1,72407755 | 0,01912303 |
| **Retinol metabolism** | L vs P | -1,69770256 | 0,02143111 |
| **Th1 and Th2 cell differentiation** | **L vs P** | **1,68189487** | **0,02143111** |
| Gastric acid secretion | L vs P | 1,63419438 | 0,02483535 |
| Valine, leucine and isoleucine degradation | L vs P | 1,7353794 | 0,02561097 |
| Primary immunodeficiency | L vs P | 1,83028683 | 0,02609136 |
| Leukocyte transendothelial migration | L vs P | 1,59447437 | 0,02719622 |
| Drug metabolism - other enzymes | L vs P | -1,61232898 | 0,02761425 |
| Estrogen signaling pathway | L vs P | 1,55208501 | 0,03013447 |
| Hepatitis C | L vs P | 1,5037 | 0,03134674 |
| alpha-Linolenic acid metabolism | L vs P | 1,70151343 | 0,03452456 |
| **Th17 cell differentiation** | **L vs P** | **1,60396027** | **0,03526215** |
| Biosynthesis of cofactors | L vs P | -1,36333205 | 0,03526215 |
| Fatty acid biosynthesis | L vs P | 1,67845253 | 0,04011674 |
| Butanoate metabolism | L vs P | 1,76610235 | 0,04046589 |
| Bladder cancer | L vs P | 1,61637916 | 0,04046589 |
| Amyotrophic lateral sclerosis | L vs P | 1,33354184 | 0,04066255 |
| Mismatch repair | L vs P | -1,73454391 | 0,04641848 |
| Staphylococcus aureus infection | L vs NL | -2,54932554 | 7,6379E-07 |
| **Cell cycle** | **L vs NL** | **2,16516678** | **7,6379E-07** |
| **Tyrosine metabolism** | **L vs NL** | **-2,32194355** | **0,00013242** |
| Intestinal immune network for IgA production | L vs NL | -2,27521529 | 0,00017033 |
| Rheumatoid arthritis | L vs NL | -2,23581081 | 0,00017947 |
| Malaria | L vs NL | -2,35252758 | 0,00018653 |
| Cell adhesion molecules | L vs NL | -2,09009499 | 0,00033184 |
| Salivary secretion | L vs NL | -2,17450627 | 0,00065633 |
| Complement and coagulation cascades | L vs NL | -2,19354355 | 0,0012977 |
| **Melanogenesis** | **L vs NL** | **-2,03036552** | **0,0012977** |
| **PPAR signaling pathway** | **L vs NL** | **-2,13113519** | **0,00154728** |
| African trypanosomiasis | L vs NL | -2,19249443 | 0,00243267 |
| Hematopoietic cell lineage | L vs NL | -2,03135757 | 0,00369709 |
| Asthma | L vs NL | -2,03610259 | 0,00574019 |
| Systemic lupus erythematosus | L vs NL | -1,99179168 | 0,00705385 |
| Tuberculosis | L vs NL | -1,78936246 | 0,00705385 |
| Mismatch repair | L vs NL | 2,08323902 | 0,00705385 |
| Viral myocarditis | L vs NL | -1,96183476 | 0,00741246 |
| Calcium signaling pathway | L vs NL | -1,75233243 | 0,00760544 |
| Nucleocytoplasmic transport | L vs NL | 1,72900371 | 0,00760544 |
| Ribosome | L vs NL | 1,63726463 | 0,00801718 |
| Dilated cardiomyopathy | L vs NL | -1,86253526 | 0,00925134 |
| **Phagosome** | **L vs NL** | **-1,72509009** | **0,00940486** |
| Ascorbate and aldarate metabolism | L vs NL | 2,01716246 | 0,01091564 |

| | | | |
|---|---|---|---|
| cGMP-PKG signaling pathway | L vs NL | -1,75972322 | 0,01166118 |
| Gastric acid secretion | L vs NL | -1,84032866 | 0,01886608 |
| Pancreatic secretion | L vs NL | -1,76458595 | 0,01886608 |
| **Chemokine signaling pathway** | **L vs NL** | **-1,67758923** | **0,02212582** |
| Cardiac muscle contraction | L vs NL | -1,86642855 | 0,02258258 |
| Leukocyte transendothelial migration | L vs NL | -1,75049227 | 0,02258258 |
| Type I diabetes mellitus | L vs NL | -1,94719486 | 0,02292594 |
| DNA replication | L vs NL | 1,9068114 | 0,02770273 |
| Alcoholic liver disease | L vs NL | -1,71694643 | 0,02814018 |
| Estrogen signaling pathway | L vs NL | -1,71903858 | 0,02989151 |
| Insulin secretion | L vs NL | -1,76600127 | 0,03186936 |
| Type II diabetes mellitus | L vs NL | -1,89752361 | 0,03384114 |
| Autoimmune thyroid disease | L vs NL | -1,93250157 | 0,03693405 |
| Allograft rejection | L vs NL | -1,93250157 | 0,03693405 |
| Graft-versus-host disease | L vs NL | -1,90474002 | 0,03693405 |
| Vascular smooth muscle contraction | L vs NL | -1,68030335 | 0,03693405 |
| Carbohydrate digestion and absorption | L vs NL | -1,77092519 | 0,03731056 |
| **Th17 cell differentiation** | **L vs NL** | **-1,74347786** | **0,03731056** |
| **Antigen processing and presentation** | **L vs NL** | **-1,72226817** | **0,03731056** |
| Hypertrophic cardiomyopathy | L vs NL | -1,72193704 | 0,03731056 |
| Pertussis | L vs NL | -1,72010768 | 0,03731056 |
| Circadian entrainment | L vs NL | -1,71203316 | 0,03731056 |
| Arrhythmogenic right ventricular cardiomyopathy | L vs NL | -1,70352699 | 0,03731056 |
| **Natural killer cell mediated cytotoxicity** | **L vs NL** | **-1,66681409** | **0,03731056** |
| Cytoskeleton in muscle cells | L vs NL | -1,56096598 | 0,03731056 |
| Diabetic cardiomyopathy | L vs NL | -1,53651312 | 0,03731056 |
| Epstein-Barr virus infection | L vs NL | -1,53532078 | 0,03731056 |
| Herpes simplex virus 1 infection | L vs NL | -1,57205292 | 0,03839106 |
| Platelet activation | L vs NL | -1,65121919 | 0,04108885 |
| Lysosome | L vs NL | -1,58097567 | 0,04433027 |
| Ribosome | P vs NL | 2,43453081 | 1,809E-12 |
| **Oxidative phosphorylation** | **P vs NL** | **2,36718206** | **1,627E-09** |
| Chemical carcinogenesis - reactive oxygen species | P vs NL | 2,12567118 | 3,1775E-08 |
| Amyotrophic lateral sclerosis | P vs NL | 1,95753736 | 5,6528E-08 |
| Thermogenesis | P vs NL | 2,0166658 | 2,1021E-07 |
| Diabetic cardiomyopathy | P vs NL | 2,02493929 | 3,2749E-07 |
| Parkinson disease | P vs NL | 1,98274573 | 3,2749E-07 |
| Huntington disease | P vs NL | 1,92547748 | 3,2749E-07 |
| Prion disease | P vs NL | 1,93375355 | 1,3093E-06 |
| **Peroxisome** | **P vs NL** | **2,22609124** | **2,67E-06** |
| Carbon metabolism | P vs NL | 2,12410465 | 9,4118E-06 |
| Non-alcoholic fatty liver disease | P vs NL | 1,97899843 | 1,0534E-05 |
| Alzheimer disease | P vs NL | 1,7215675 | 4,7179E-05 |
| Cytoskeleton in muscle cells | P vs NL | -1,90235145 | 5,1755E-05 |
| **Fatty acid metabolism** | **P vs NL** | **2,11066585** | **9,9675E-05** |
| Calcium signaling pathway | P vs NL | -1,94510045 | 0,00010791 |

| | | | |
|---|---|---|---|
| Dilated cardiomyopathy | P vs NL | -2,06708309 | 0,00064934 |
| Pathways of neurodegeneration - multiple diseases | P vs NL | 1,56163799 | 0,00069777 |
| Pyruvate metabolism | P vs NL | 2,1016309 | 0,00073701 |
| Citrate cycle (TCA cycle) | P vs NL | 2,07027601 | 0,00088888 |
| African trypanosomiasis | P vs NL | -2,23022842 | 0,00139616 |
| Fatty acid elongation | P vs NL | 2,04636459 | 0,00139616 |
| Vascular smooth muscle contraction | P vs NL | -1,8721075 | 0,00154846 |
| Hypertrophic cardiomyopathy | P vs NL | -1,80758059 | 0,00637617 |
| Oxytocin signaling pathway | P vs NL | -1,67996673 | 0,00855714 |
| Butanoate metabolism | P vs NL | 1,94341977 | 0,00949526 |
| Terpenoid backbone biosynthesis | P vs NL | 1,88697514 | 0,01147558 |
| Insulin secretion | P vs NL | -1,84941055 | 0,01162878 |
| Hormone signaling | P vs NL | -1,69677889 | 0,01229754 |
| Biosynthesis of unsaturated fatty acids | P vs NL | 1,86806895 | 0,01293322 |
| Biosynthesis of amino acids | P vs NL | 1,78379035 | 0,01370503 |
| **Tyrosine metabolism** | **P vs NL** | **-2,01722223** | **0,01397578** |
| Steroid biosynthesis | P vs NL | 1,91419391 | 0,01397578 |
| Platelet activation | P vs NL | -1,62666466 | 0,0145248 |
| Fatty acid degradation | P vs NL | 1,85771394 | 0,01527478 |
| Mucin type O-glycan biosynthesis | P vs NL | 1,83751527 | 0,01527478 |
| **Cell cycle** | **P vs NL** | **1,53983711** | **0,0203955** |
| Glyoxylate and dicarboxylate metabolism | P vs NL | 1,78708435 | 0,0204455 |
| Spliceosome | P vs NL | -1,55229394 | 0,02295253 |
| Retrograde endocannabinoid signaling | P vs NL | 1,54173678 | 0,02295253 |
| 2-Oxocarboxylic acid metabolism | P vs NL | 1,75033825 | 0,02316649 |
| Bile secretion | P vs NL | -1,78271462 | 0,02574062 |
| Malaria | P vs NL | -1,81912452 | 0,02741936 |
| Motor proteins | P vs NL | -1,59502542 | 0,03459158 |
| cGMP-PKG signaling pathway | P vs NL | -1,56919031 | 0,03508619 |
| Proteasome | P vs NL | 1,68561141 | 0,03594641 |
| Adrenergic signaling in cardiomyocytes | P vs NL | -1,56573783 | 0,04185303 |

**Table S4.** Connectivity Map analysis outcomes ranked on raw connectivity scores (raw_cs), filtered for the A375 cell line, perturbagen type “trt_cp,” and high significance (fdr_q_nlog10 ≥ 15.35). Compounds discussed in the manuscript are shown in bold.

| Compound | Cell line | Treatment code | raw_cs | fdr_q_nlog10 |
|---|---|---|---|---|
| BRD-K07689498 | A375 | trt_cp | -0.33 | 15.35 |
| RS-23597-190 | A375 | trt_cp | -0.33 | 15.35 |
| BRD-K16947188 | A375 | trt_cp | -0.33 | 15.35 |
| BRD-K19642661 | A375 | trt_cp | -0.33 | 15.35 |
| BRD-K20834925 | A375 | trt_cp | -0.33 | 15.35 |
| BRD-A21406558 | A375 | trt_cp | -0.33 | 15.35 |
| BMS-265246 | A375 | trt_cp | -0.33 | 15.35 |
| UCL-2077 | A375 | trt_cp | -0.33 | 15.35 |
| nitrendipine | A375 | trt_cp | -0.33 | 15.35 |

| | | | | |
|---|---|---|---|---|
| ceritinib | A375 | trt_cp | -0.33 | 15.35 |
| BRD-K76535108 | A375 | trt_cp | -0.33 | 15.35 |
| BRD-A64850503 | A375 | trt_cp | -0.33 | 15.35 |
| SA-247714 | A375 | trt_cp | -0.34 | 15.35 |
| BRD-K95638669 | A375 | trt_cp | -0.34 | 15.35 |
| BRD-K05814022 | A375 | trt_cp | -0.34 | 15.35 |
| **dasatinib** | **A375** | **trt_cp** | **-0.34** | **15.35** |
| BRD-K94390040 | A375 | trt_cp | -0.34 | 15.35 |
| BRD-K72753750 | A375 | trt_cp | -0.34 | 15.35 |
| **PF-543** | **A375** | **trt_cp** | **-0.34** | **15.35** |
| BRD-K38672372 | A375 | trt_cp | -0.34 | 15.35 |
| promazine | A375 | trt_cp | -0.34 | 15.35 |
| BRD-K86108784 | A375 | trt_cp | -0.34 | 15.35 |
| **PRT-062607** | **A375** | **trt_cp** | **-0.34** | **15.35** |
| SB-271046 | A375 | trt_cp | -0.34 | 15.35 |
| WZ-4-145 | A375 | trt_cp | -0.34 | 15.35 |
| BRD-K84571636 | A375 | trt_cp | -0.34 | 15.35 |
| berbamine | A375 | trt_cp | -0.34 | 15.35 |
| PD-168393 | A375 | trt_cp | -0.34 | 15.35 |
| **hydrocortisone** | **A375** | **trt_cp** | **-0.34** | **15.35** |
| BRD-K55633328 | A375 | trt_cp | -0.34 | 15.35 |
| BRD-K70663894 | A375 | trt_cp | -0.34 | 15.35 |
| LY-2835219 | A375 | trt_cp | -0.34 | 15.35 |
| BRD-K51267267 | A375 | trt_cp | -0.34 | 15.35 |
| toremifene | A375 | trt_cp | -0.34 | 15.35 |
| demecarium | A375 | trt_cp | -0.34 | 15.35 |
| regorafenib | A375 | trt_cp | -0.34 | 15.35 |
| metixene | A375 | trt_cp | -0.34 | 15.35 |
| vandetanib | A375 | trt_cp | -0.34 | 15.35 |
| CCT-031374 | A375 | trt_cp | -0.34 | 15.35 |
| VU-0155056 | A375 | trt_cp | -0.34 | 15.35 |
| ZK-164015 | A375 | trt_cp | -0.34 | 15.35 |
| BRD-K27630941 | A375 | trt_cp | -0.34 | 15.35 |
| picrotoxin | A375 | trt_cp | -0.34 | 15.35 |
| BRD-K07005393 | A375 | trt_cp | -0.34 | 15.35 |
| BRD-K87779450 | A375 | trt_cp | -0.34 | 15.35 |
| pranlukast | A375 | trt_cp | -0.34 | 15.35 |
| etoposide | A375 | trt_cp | -0.34 | 15.35 |
| hesperidin | A375 | trt_cp | -0.34 | 15.35 |
| DBeQ | A375 | trt_cp | -0.34 | 15.35 |
| JAS07-008 | A375 | trt_cp | -0.34 | 15.35 |
| phenamil | A375 | trt_cp | -0.34 | 15.35 |
| KIN001-266 | A375 | trt_cp | -0.34 | 15.35 |
| TAK-875 | A375 | trt_cp | -0.35 | 15.35 |
| BRD-K84770158 | A375 | trt_cp | -0.35 | 15.35 |
| BRD-A76641868 | A375 | trt_cp | -0.35 | 15.35 |

| | | | | |
|---|---|---|---|---|
| BRD-K71599932 | A375 | trt_cp | -0.35 | 15.35 |
| SB-205607 | A375 | trt_cp | -0.35 | 15.35 |
| NVP-ADW742 | A375 | trt_cp | -0.35 | 15.35 |
| **lapatinib** | **A375** | **trt_cp** | **-0.35** | **15.35** |
| BRD-K26840329 | A375 | trt_cp | -0.35 | 15.35 |
| **methotrexate** | **A375** | **trt_cp** | **-0.35** | **15.35** |
| BRD-K55210983 | A375 | trt_cp | -0.35 | 15.35 |
| BRD-K56731825 | A375 | trt_cp | -0.35 | 15.65 |
| RG-13022 | A375 | trt_cp | -0.35 | 15.65 |
| JDTic | A375 | trt_cp | -0.35 | 15.65 |
| ST-4042556 | A375 | trt_cp | -0.35 | 15.65 |
| G-5555 | A375 | trt_cp | -0.35 | 15.65 |
| SCH-772984 | A375 | trt_cp | -0.35 | 15.65 |
| SA-1921867 | A375 | trt_cp | -0.35 | 15.65 |
| camicinal | A375 | trt_cp | -0.35 | 15.65 |
| BRD-K58493165 | A375 | trt_cp | -0.35 | 15.65 |
| PSB-36 | A375 | trt_cp | -0.35 | 15.65 |
| ABT-239 | A375 | trt_cp | -0.35 | 15.65 |
| niguldipine | A375 | trt_cp | -0.35 | 15.65 |
| carmofur | A375 | trt_cp | -0.35 | 15.65 |
| **ibrutinib** | **A375** | **trt_cp** | **-0.35** | **15.65** |
| bendamustine | A375 | trt_cp | -0.35 | 15.65 |
| **ARRY-334543** | **A375** | **trt_cp** | **-0.35** | **15.65** |
| BRD-K74461819 | A375 | trt_cp | -0.36 | 15.65 |
| BRD-K83834119 | A375 | trt_cp | -0.36 | 15.65 |
| vortioxetine | A375 | trt_cp | -0.36 | 15.65 |
| bazedoxifene | A375 | trt_cp | -0.36 | 15.65 |
| tariquidar | A375 | trt_cp | -0.36 | 15.65 |
| protriptyline | A375 | trt_cp | -0.36 | 15.65 |
| raloxifene | A375 | trt_cp | -0.36 | 15.65 |
| BRD-K93158953 | A375 | trt_cp | -0.36 | 15.65 |
| **medrysone** | **A375** | **trt_cp** | **-0.36** | **15.65** |
| BRD-K46065939 | A375 | trt_cp | -0.36 | 15.65 |
| BRD-K44573794 | A375 | trt_cp | -0.36 | 15.65 |
| prochlorperazine | A375 | trt_cp | -0.36 | 15.65 |
| zaldaride | A375 | trt_cp | -0.36 | 15.65 |
| **BIX-01294** | **A375** | **trt_cp** | **-0.36** | **15.65** |
| BRD-K71670746 | A375 | trt_cp | -0.36 | 15.65 |
| CAM-9-027-3 | A375 | trt_cp | -0.36 | 15.65 |
| BRD-K67662618 | A375 | trt_cp | -0.36 | 15.65 |
| brequinar | A375 | trt_cp | -0.36 | 15.65 |
| loperamide | A375 | trt_cp | -0.36 | 15.65 |
| BRD-A72575012 | A375 | trt_cp | -0.37 | 15.65 |
| clomifene | A375 | trt_cp | -0.37 | 15.65 |
| cepharanthine | A375 | trt_cp | -0.37 | 15.65 |
| ST-014075 | A375 | trt_cp | -0.37 | 15.65 |

| | | | | |
|---|---|---|---|---|
| **STA-5326** | **A375** | **trt_cp** | **-0.37** | **15.65** |
| clomifene | A375 | trt_cp | -0.37 | 15.65 |
| BMS-833923 | A375 | trt_cp | -0.37 | 15.65 |
| serdemetan | A375 | trt_cp | -0.37 | 15.65 |
| trifluoperazine | A375 | trt_cp | -0.37 | 15.65 |
| BRD-K20492338 | A375 | trt_cp | -0.37 | 15.65 |
| thiothixene | A375 | trt_cp | -0.37 | 15.65 |
| BRD-K87367419 | A375 | trt_cp | -0.37 | 15.65 |
| BRD-K60084530 | A375 | trt_cp | -0.37 | 15.65 |
| BRD-K80954687 | A375 | trt_cp | -0.37 | 15.65 |
| BRD-K06816133 | A375 | trt_cp | -0.38 | 15.65 |
| BRD-K75708657 | A375 | trt_cp | -0.38 | 15.65 |
| **dexamethasone** | **A375** | **trt_cp** | **-0.38** | **15.65** |
| XMD-1499 | A375 | trt_cp | -0.38 | 15.65 |
| BRD-K89824424 | A375 | trt_cp | -0.38 | 15.65 |
| **GSK-269962** | **A375** | **trt_cp** | **-0.38** | **15.65** |
| BRD-K63158888 | A375 | trt_cp | -0.38 | 15.65 |
| siramesine | A375 | trt_cp | -0.38 | 15.65 |
| **NVP-BSK805** | **A375** | **trt_cp** | **-0.38** | **15.65** |
| flumatinib | A375 | trt_cp | -0.38 | 15.65 |
| **GSK-343** | **A375** | **trt_cp** | **-0.38** | **15.65** |
| BRD-K80786583 | A375 | trt_cp | -0.38 | 15.65 |
| SA-1921085 | A375 | trt_cp | -0.39 | 15.65 |
| BRD-K87125912 | A375 | trt_cp | -0.39 | 15.65 |
| BRD-K97194822 | A375 | trt_cp | -0.39 | 15.65 |
| **CGP-53353** | **A375** | **trt_cp** | **-0.39** | **15.65** |
| BRD-K92543914 | A375 | trt_cp | -0.39 | 15.65 |
| **dipyridamole** | **A375** | **trt_cp** | **-0.39** | **15.65** |
| BRD-K93950432 | A375 | trt_cp | -0.39 | 15.65 |
| BRD-K80779790 | A375 | trt_cp | -0.39 | 15.65 |
| metergoline | A375 | trt_cp | -0.39 | 15.65 |
| BRD-K03006541 | A375 | trt_cp | -0.39 | 15.65 |
| dronedarone | A375 | trt_cp | -0.39 | 15.65 |
| amodiaquine | A375 | trt_cp | -0.39 | 15.65 |
| BAFILOMYCIN-A1 | A375 | trt_cp | -0.39 | 15.65 |
| midodrine | A375 | trt_cp | -0.40 | 15.65 |
| BRD-K06956503 | A375 | trt_cp | -0.40 | 15.65 |
| **UNC-1999** | **A375** | **trt_cp** | **-0.40** | **15.65** |
| mibefradil | A375 | trt_cp | -0.40 | 15.65 |
| BRD-K31029552 | A375 | trt_cp | -0.40 | 15.65 |
| sertindole | A375 | trt_cp | -0.40 | 15.65 |
| SA-1919584 | A375 | trt_cp | -0.41 | 15.65 |
| BRD-K25379780 | A375 | trt_cp | -0.41 | 15.65 |
| DDR1-IN-1 | A375 | trt_cp | -0.41 | 15.65 |
| RS-39604 | A375 | trt_cp | -0.41 | 15.65 |
| CAY-10594 | A375 | trt_cp | -0.41 | 15.65 |

| | | | | |
|---|---|---|---|---|
| N-benzylnaltrindole | A375 | trt_cp | -0.41 | 15.65 |
| BRD-K49477330 | A375 | trt_cp | -0.41 | 15.65 |
| amiodarone | A375 | trt_cp | -0.41 | 15.65 |
| DL-PDMP | A375 | trt_cp | -0.41 | 15.65 |
| fluphenazine | A375 | trt_cp | -0.41 | 15.65 |
| maprotiline | A375 | trt_cp | -0.41 | 15.65 |
| thioridazine | A375 | trt_cp | -0.42 | 15.65 |
| **SU-11274** | **A375** | **trt_cp** | **-0.42** | **15.65** |
| amitriptyline | A375 | trt_cp | -0.42 | 15.65 |
| SGI-1776 | A375 | trt_cp | -0.43 | 15.65 |
| BRD-K41335306 | A375 | trt_cp | -0.43 | 15.65 |
| BRD-K67161258 | A375 | trt_cp | -0.43 | 15.65 |
| BRD-K15050703 | A375 | trt_cp | -0.43 | 15.65 |
| **golvatinib** | **A375** | **trt_cp** | **-0.43** | **15.65** |
| **cediranib** | **A375** | **trt_cp** | **-0.44** | **15.65** |
| NNC-05-2090 | A375 | trt_cp | -0.44 | 15.65 |
| BRD-K59333713 | A375 | trt_cp | -0.44 | 15.65 |
| BRD-K13659644 | A375 | trt_cp | -0.44 | 15.65 |
| **LLY-507** | **A375** | **trt_cp** | **-0.44** | **15.65** |
| BRD-K91663486 | A375 | trt_cp | -0.46 | 15.65 |
| terconazole | A375 | trt_cp | -0.47 | 15.65 |
| SA-1921456 | A375 | trt_cp | -0.47 | 15.65 |
| trimipramine | A375 | trt_cp | -0.47 | 15.65 |
